# Seroprevalence of anti-SARS-CoV-2 IgG antibodies, risk factors for infection and associated symptoms in Geneva, Switzerland: a population-based study

**DOI:** 10.1101/2020.12.16.20248180

**Authors:** Aude Richard, Ania Wisniak, Javier Perez-Saez, Henri Garrison-Desany, Dusan Petrovic, Giovanni Piumatti, Hélène Baysson, Attilio Picazio, Francesco Pennacchio, David De Ridder, François Chappuis, Nicolas Vuilleumier, Nicola Low, Samia Hurst, Isabella Eckerle, Antoine Flahault, Laurent Kaiser, Andrew S Azman, Idris Guessous, Silvia Stringhini, For the SEROCOV-POP study group

## Abstract

**Background:** Population-based serological surveys provide a means for assessing the immunologic landscape of a community, without the biases related to health-seeking behaviors and testing practices typically associated with rt-PCR testing. This study assesses SARS-CoV-2 seroprevalence over the first epidemic wave in Canton Geneva, Switzerland, as well as biological and socio-economic risk factors for infection and symptoms associated with IgG seropositivity.

**Methods and findings:** Between April 6 and June 30, 2020, former participants of a yearly representative cross-sectional survey of the 20-75-year-old population of the canton of Geneva were invited to participate in a seroprevalence study, along with household members five years and older. We collected blood and tested it for anti-SARS-CoV-2 immunoglobulins G (IgG). Questionnaires were self-administered. We estimated seroprevalence with a Bayesian model accounting for test performance and sampling design. We included 8344 participants (53.5% women, mean age 46.9 years). The population-level seroprevalence over the 12-week study period was 7.8 % (95% Credible Interval (CrI) 6.8-8.9), accounting for sex, age and household random effects. Seroprevalence was highest among 18-49 year olds (9.5%, 95%CrI 8.1-10.9), with young children (5-9 years) and those >65 years having significantly lower seroprevalence (4.3% and 4.7-5.4% respectively). Men were more likely to be seropositive than women (relative risk 1.2, 95%CrI 1.1-1.4). Odds of seropositivity were reduced for female retirees (0.46, 95%CI 0.23-0.93) and unemployed men (0.35, 95%CI 0.13-1.0) compared to employed individuals, and for current smokers (0.36, 95%CI 0.23-0.55) compared to never-smokers. We found no significant association between occupation, level of education, neighborhood income and the risk of being seropositive. Symptoms most strongly associated with seropositivity were anosmia/dysgeusia, loss of appetite, fever, fatigue and myalgia and/or arthralgia. Thirteen percent of seropositive participants reported no symptoms.

**Conclusions:** Our results confirm a low population seroprevalence of anti-SARS-CoV-2 antibodies after the first wave in Geneva, a region hard hit by the COVID-19 pandemic. Socioeconomic factors were not associated with seropositivity in this sample. The elderly and young children were less frequently seropositive, though it is not clear how biology and behaviors shape these differences. These specificities should be considered when assessing the need for targeted public health measures.

## Introduction

Seroprevalence studies around the world have established that only a small proportion of the population had been infected by SARS-CoV-2 towards the end of the first wave in spring 2020 with most seroprevalence estimates ranging from <0.1 to more than 20%, depending on the setting and targeted populations.(1–3) Preliminary results from the first five weeks of our 12-week population-based serosurvey conducted in the canton of Geneva, Switzerland, in April and May, showed a weekly seroprevalence between 4.8% and 10.9%.(4) Other areas with similar clinical incidence rates, such as certain regions in France, Italy and Spain showed similar results,(5–7) while the least affected regions in Spanish and French national surveys showed seroprevalences of less than 3%.(5–8)

Socio-economic risk factors for COVID-19 such as low income, social deprivation and overcrowded living conditions have been identified in several studies based on molecular testing in the United Kingdom, the United States, and Brazil.(9–13) However, some of those studies included only hospitalized patients or individuals with severe COVID-19. Data on confirmed COVID-19 cases are influenced by testing policies and accessibility and comprise only a minority of the whole infected population. Serosurveys have the potential to assess the real extent of an outbreak in a given geographical region and provide information on risk factors, transmission and infection fatality rate without relying on clinical surveillance and virologic confirmation. Nevertheless, only a few population-based serostudies assessed biological and socio-economic risk factors for seropositivity. These have generally highlighted significant relationships between Black, Asian and Hispanic ethnicity, adult age (varying across studies), living in a deprived or dense area and large household size, and the risk of being seropositive.(5–8,14,15) While working in the healthcare field or essential services has been shown to be associated with a higher risk of seropositivity, other individual factors such as education level and occupational category are rarely described.(6,7,14,16) Serosurveys can also provide a means to quantify the full spectrum of clinical symptoms associated with SARS-CoV-2, including mild and asymptomatic infections, which are often missed in clinical surveillance.

In Switzerland, the first cases of COVID-19 were recorded at the end of February 2020, followed by a first epidemic wave that took place during March and April 2020.(17) Strict non-pharmaceutical measures started on March 16^th^ and were progressively relaxed from April 27^th^ 2020 when a total of 29’313 cases and 427 deaths had been reported in the country.(18) In this study, we describe the age-specific seroprevalence of anti-SARS-CoV-2 antibodies between April and June 2020 in Geneva, Switzerland, and examine risk factors for and symptoms associated with seropositivity.

## Methods

### Study design and participants

We recruited participants between April 6^th^ and June 30^th^, 2020. The recruitment strategy and serological data collection have previously been described in detail.(4) Briefly, SEROCoV-POP is a population-based observational seroprevalence study. Participants were invited progressively between April 2^nd^ and June 9^th^, by email and postal mail, from the study population of a yearly health survey called Bus Santé,(19) which is representative of the population of canton Geneva aged between 20 and 74 years old. Participants enrolled in the SEROCoV-POP study were invited to bring members of their household aged 5 years and older. Each person participated only once (one round). Of note, during the study period, the country was initially under lockdown, then, measures were progressively relaxed in three consecutive periods.(20) Participants provided written informed consent to participate in the study, and parents or legal guardians gave consent for children under 18 years. The study protocol was approved by the Cantonal Research Ethics Commission of Geneva, Switzerland (CER16-363) and is available online at https://static1.squarespace.com/static/5e7dd8f02d3bc353fbb05121/t/5e887016c7fa18312c3e00fa/1585999900784/Protocole_SEROCOV-POP30.03.pdf.

### Laboratory analysis

Anti-SARS-CoV-2 immunoglobulin G (IgG) serological status was assessed using a commercially available enzyme-linked immunosorbent assay (ELISA) (Euroimmun; Lübeck, Germany #EI 2606-9601 G) targeting the S1 domain of the spike protein. We used the manufacturer’s recommended cutoff for positivity (≥1·1) with a reported sensitivity of 93% and a specificity of 100% estimated in a diagnostic validation study conducted in the same lab as these analyses.(21) We tested all serum with an ELISA IgG ratio of ≥ 0.5 with an immunofluorescence assay (IFA) and classified these individuals with results from this test (when performed) in sensitivity analyses.

### Survey questions and coding

Participants could choose to fill in the questionnaire online at home or complete it at the study site on a tablet computer or on paper.

Questionnaires included questions about socio-demographic characteristics, symptoms, exposure to SARS-CoV-2-infected individuals and preventive behaviors in relation to the epidemic. Socio-demographic variables included age, sex, employment status, professional occupation, professional changes due to the COVID-19 pandemic, and educational level. Educational level was based on the national Swiss system, with “mandatory education” applying to all children ages 4 to 14, “secondary education” corresponding to high school (15 to 18), and “university” corresponding to undergraduate and postgraduate studies.

Participants were also asked to self-identify their occupational sector and profession (the latter was entered as free text). We mapped this information using the European Socio-Economic Classifications (ESEC). From the 10 levels of classification, in order to maintain power, this was reduced to 8 categories: (1) professional/manager (ESEC 1-2), (2) higher grade white-collar workers (ESEC 2) (3) independent workers (ESEC 4-5), (4) lower grade white-collar workers(ESEC 7), (5) blue-collar, skilled, semi-skilled and non-skilled workers (ESEC 6, 8, 9), (6) other full-time workers to whom the ESEC categories were not applicable; as well as from ESEC class 10: (7) students or retirees, and (8) unemployed.

Participants were asked to self-report all symptoms not related to a known chronic condition experienced since January 2020, and to specify if these appeared as part of one or several distinct episodes. We defined participants as asymptomatic if they didn’t report any symptoms. Data were also collected on chronic health conditions, smoking status and height and weight for body mass index (BMI) calculation. The full study questionnaire is provided in the Supplement.

### Other data sources

Data on individual-level income was not collected in the questionnaire. However, based on participants’ neighborhood of residence, we used data from the Canton of Geneva to determine the median income for a single individual residing in participants’ neighborhoods. We grouped gross income into three categories defined as <37,000 CHF/year (70% of the median income), between 37,000 and 68,000 CHF/year, and >68,000 CHF/year (∼130% of the median income).(22)

### Statistical analysis

We estimated the overall 12-week seroprevalence using a Bayesian logistic regression model accounting for test performance, sex, age as well as within household infection clustering as previously described.(4) In our primary analyses, we used IgG ELISA results only to classify seropositivity, and performed post-stratification to obtain population-wide seroprevalence as well as by age and sex classes. As a sensitivity analysis, we used the IFA results for all participants tested with this assay. Models were implemented in the Stan probabilistic programming language. Details are provided in the Supplementary Material. Weekly crude proportions of seropositivity with binomial confidence intervals (CI) were also calculated.

For socio-economic and lifestyle risk factors, we limited our analysis to participants ≥18 years of age, resulting in a final analytic sample of 7442 participants. To test for association between socio-demographic, lifestyle and health-related risk factors and serological status, we used mixed effect logistic models including a household-level random effect, except for the area-level income analysis. To understand the sensitivity of our results to missing data, we conducted multi-level multiple imputation by chained equations using 10 datasets with 30 iterations across the dataset and reran our primary analyses.

Chi square tests were used to compare the frequencies of individual symptoms between IgG seropositive and seronegative individuals. Using logistic regression models, we estimated odds ratios (OR) and 95% CIs for IgG seropositivity according to the presence of each symptom in the overall study sample and stratified by age groups. As a sensitivity analysis, we reran the logistic regression on the individuals presenting only one symptomatic episode. To take into account the co-occurrence of symptoms, we fit multivariable logistic regression models to estimate sex-adjusted and mutually adjusted ORs. For asymptomatic status, we only adjusted the ORs for sex and age. The absence of a symptom was defined as the reference value for each symptom, while for the asymptomatic variable, not being asymptomatic was considered the reference value. Analyses were done using R statistical software.(23)

## Results

Between April 6th and June 30th, 2020, we enrolled 8’344 participants between the ages of 5 and 94 (902 children < 18 years, and 7442 adults), with a mean age of 46.9 years. The study sample included 4’379 former Bus Santé participants and 3’965 household members, corresponding to a 42% participation rate of all people invited by email (Figure S1).

Compared with the population of the Canton of Geneva, in SEROCoV-POP, the 50-74 age group was overrepresented (46.6% vs. 27.4%), while the 5-9, 20-49 and ≥75 age groups were slightly underrepresented. SEROCoV-POP participants also had generally higher levels of education than the Geneva population, with more participants having received a tertiary education (57.7% vs. 39.3%), and less having attended mandatory school only (6.8% vs. 28.1%) (Table S1).

### Seroprevalence and relative risk of seropositivity by age and sex

SARS-CoV-2 seroprevalence over the study period was 7.8 % (95% Credible Interval (CrI) 6.8-8.9, Table 1). The seroprevalence was 7.1% (95% CrI 5.5-8.7) over the first study month, increased to 9.0% (95% CrI 7.5-10.5) during the second month, before decreasing again to 7.1% (95% CrI 5.7-8.5) in the final study month (Table 1).

**Table 1.**
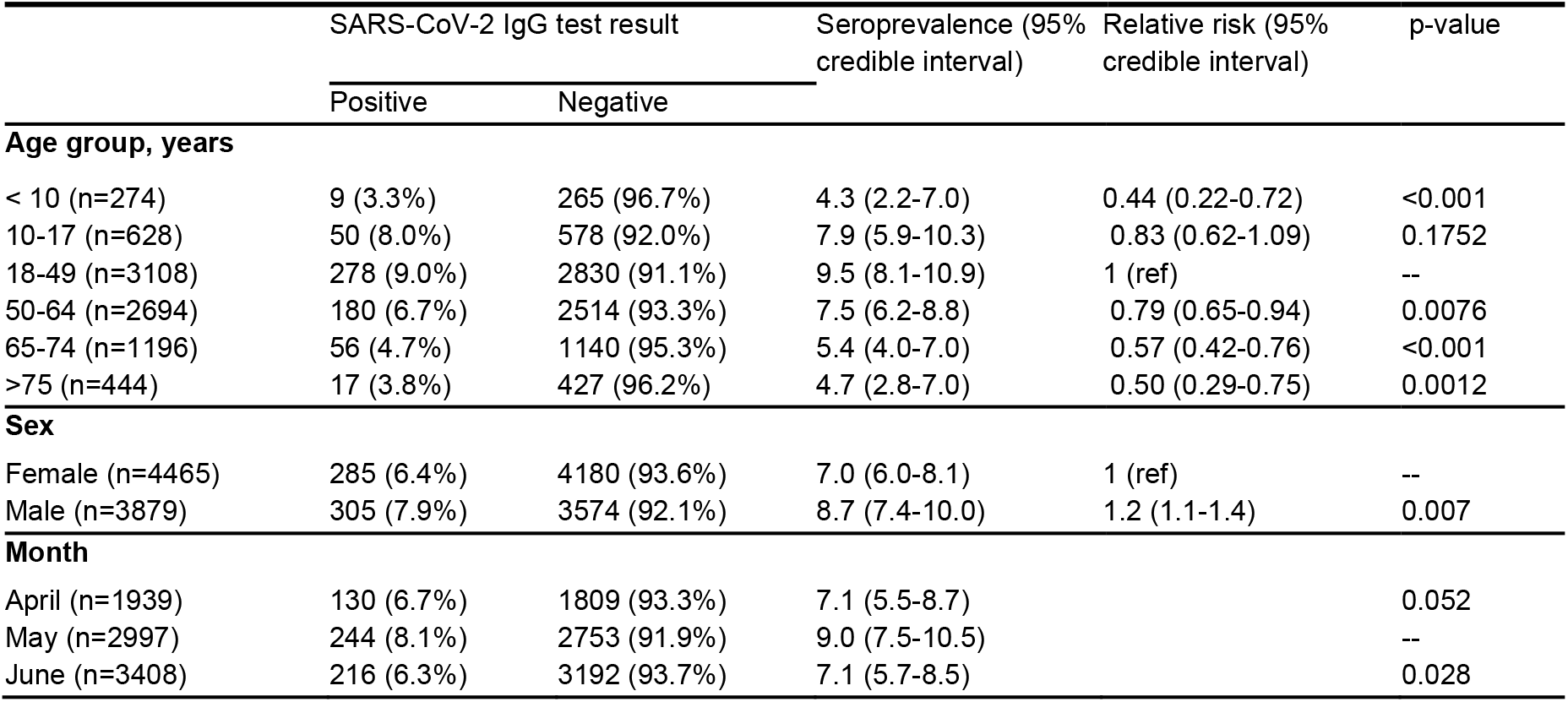
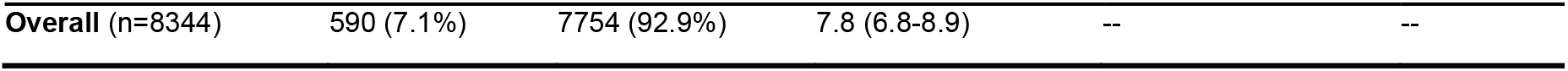
Seroprevalence estimates and relative risk of SARS-CoV-2 seropositivity (N = 8’344)

Men had a 22% (95% CrI 6-40%) higher risk of being seropositive than women. Seroprevalence was highest in the 18-49 age group (9.5%, 95% CrI: 8.1-10.9) and lowest in young children < 10 years (4.3%, 95% CrI 2.2-7.0) and the elderly ≥ 75 years (4.7%, 95% CrI 2.8-7.0). Seroprevalence estimates by 10-year age groups are provided in Table S3.

### Socio-economic, lifestyle and health-related risk factors

After adjusting for age and sex, there was a significant difference in the distribution of seropositive participants according to employment status (Table 2). The mixed effect logistic model analysis adjusting for age and sex showed lower odds of being seropositive in female retirees (OR 0.46, 95% CI 0.23-0.93) and unemployed men (OR 0.35, 95% CI 0.13-1.0) compared to employed individuals of the same sex. However, we did not observe any significant association between occupational category, educational level, area-based income and seropositivity.

**Table 2.**
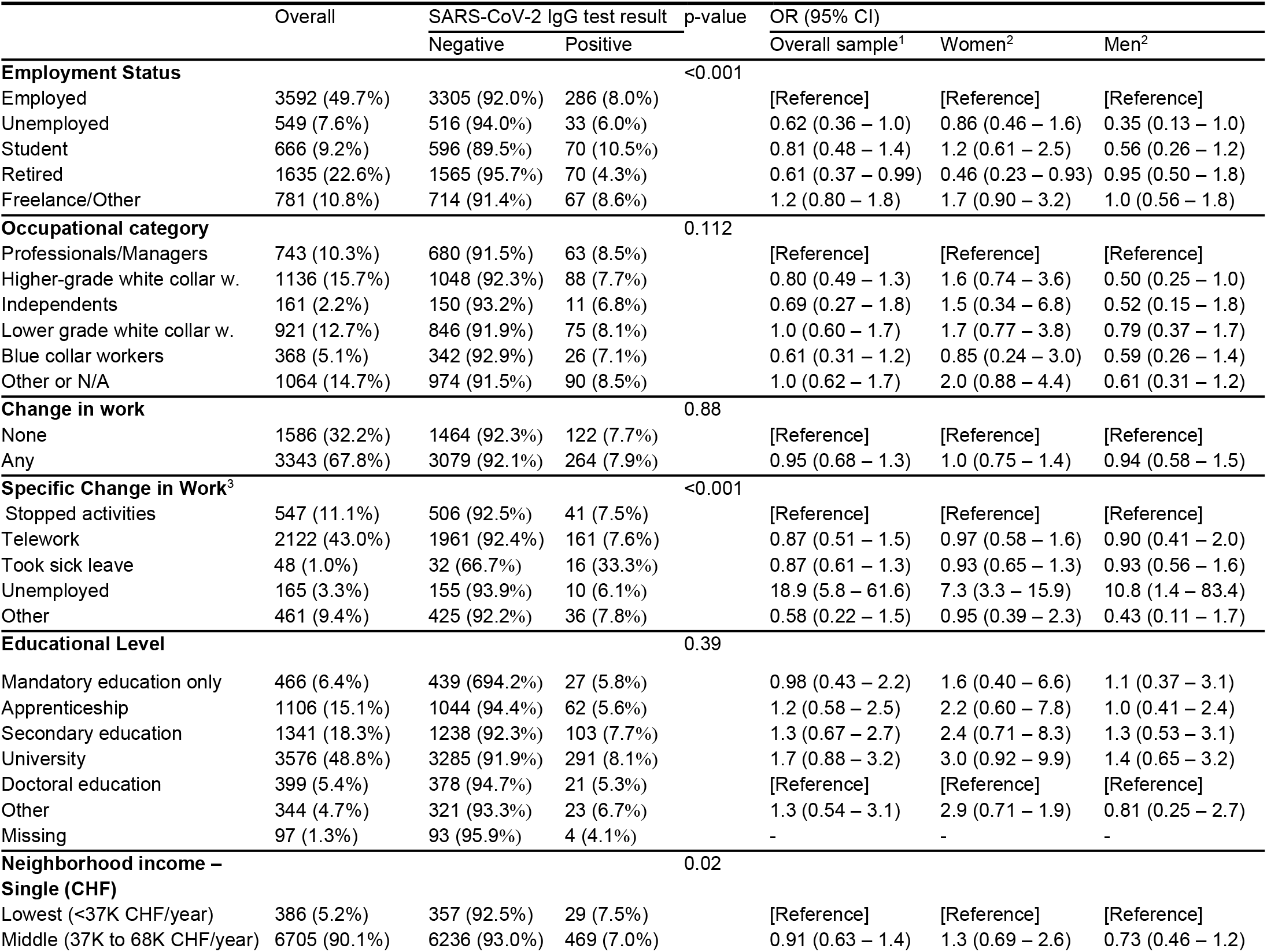
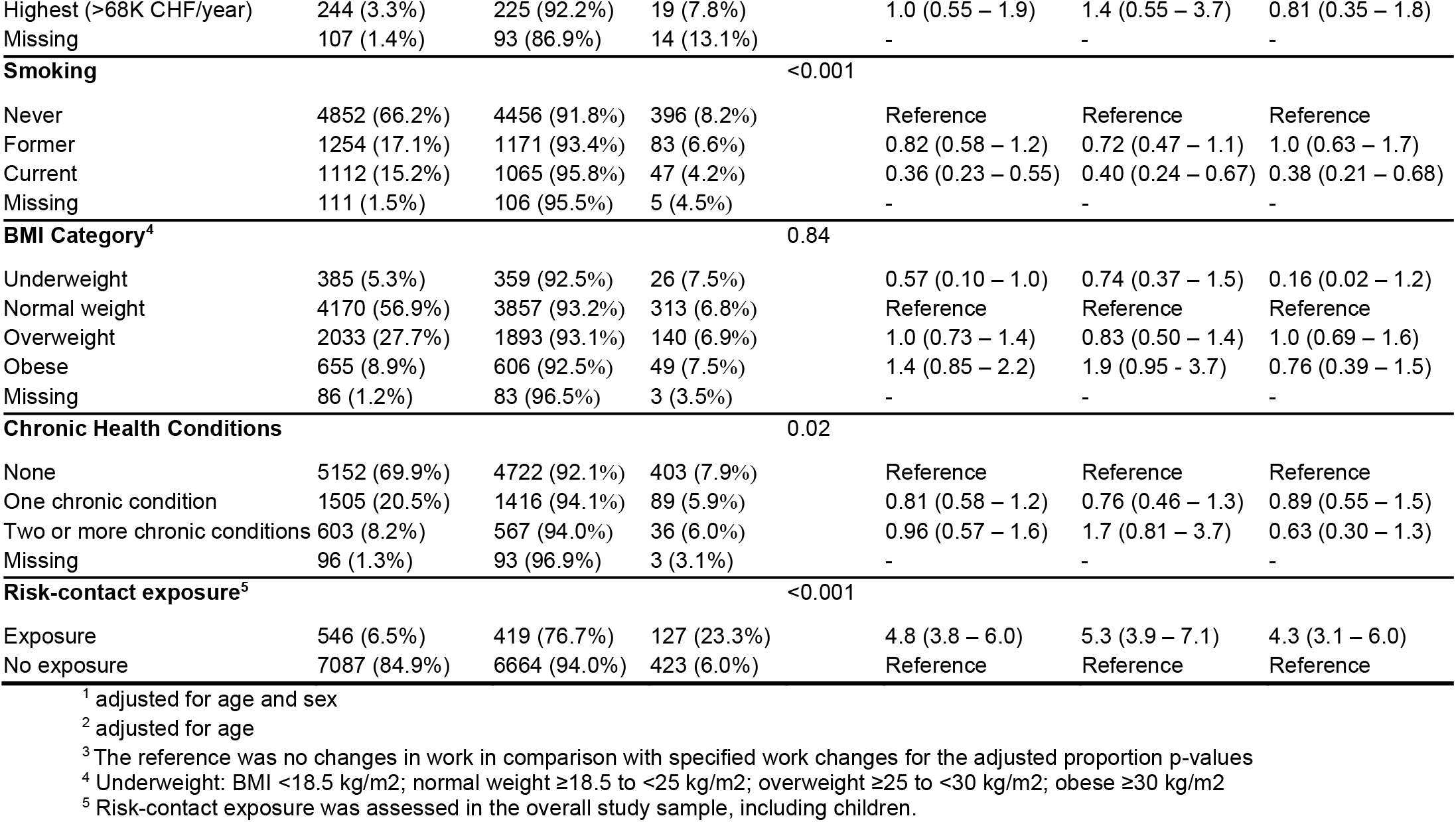
Socio-demographic, behavioral and health characteristics of the adult SEROCOV-POP study participants (N = 7’329) by SARS-CoV-2 serology test result, and association with serological status.

Overall, 67.8% of the participants reported some change in work linked to the COVID-19 epidemic, such as working remotely (43.0%), taking a sick leave (1.0%) or becoming unemployed (3.3%). A marked 33.6% of those who took a sick leave were seropositive, as opposed to 6.1% to 7.8% in all other categories.

Though not statistically significant, there seemed to be a gradient in the association of education with seropositivity, from mandatory schooling only (OR 0.98, 95% CI 0.43-2.2) all the way up to university level (OR 1.7, 95% CI 0.88-3.2), when compared to individuals with a doctoral education. This effect was stronger in women than men.

Current smokers had 64% lower odds of being seropositive compared to never smokers (OR= 0.36, 95% CI 0.23-0.55). There was no significant association between body mass index or chronic health conditions and seropositivity, although obese women tended to have higher odds of seropositivity compared to participants with a normal BMI (OR 1.9, 95% CI 0.95-3.7). A history of close contact with a confirmed SARS-CoV-2-infected person increased the odds of seropositivity nearly five-fold (OR 4.8, 95% CI 3.8-6.0) in the overall population.

### Symptoms reported over one or more episodes

The symptoms most frequently reported by seropositive participants were fatigue (57% of seropositive participants), headache (52%), sneezing and/or rhinorrhea (48%), fever (46%), cough (46%), anosmia and/or dysgeusia (44%), and myalgia and/or arthralgia (43%) (Table 3). Seronegative individuals, on the other hand, most frequently reported sneezing/rhinorrhea (32%), while only 3% reported loss of taste or smell. Thirteen percent of seropositive participants reported no symptoms, versus 43% of seronegative participants. Not reporting any symptoms during the study period reduced by five the odds of being seropositive.

**Table 3.** Frequency of symptoms reported in all age groups.

| Symptom | SARS-CoV-2 IgG test result |  | p-value |
| --- | --- | --- | --- |
|  | Positive<br>(N = 590)* | Negative<br>(N = 7,754)** |  |
| Fatigue | 332 (57%) | 1,787 (23%) | <0.001 |
| Headache | 308 (52%) | 1,936 (25%) | <0.001 |
| Sneezing/Rhinorrhea | 284 (48%) | 2,468 (32%) | <0.001 |
| Fever | 272 (46%) | 1,136 (15%) | <0.001 |
| Cough | 270 (46%) | 1,790 (23%) | <0.001 |
| Anosmia / dysgeusia | 257 (44%) | 233 (3.0%) | <0.001 |
| Myalgia / arthralgia | 252 (43%) | 1,160 (15%) | <0.001 |
| Sore throat | 200 (34%) | 1,786 (23%) | <0.001 |
| Dyspnea | 131 (22%) | 577 (7.5%) | <0.001 |
| Loss of appetite | 126 (21%) | 323 (4.2%) | <0.001 |
| Diarrhea | 121 (21%) | 749 (9.8%) | <0.001 |
| Abdominal pain | 65 (11%) | 527 (6.9%) | <0.001 |
| Nausea / vomiting | 55 (9.4%) | 374 (4.9%) | <0.001 |
| Other symptoms | 18 (3.1%) | 181 (2.4%) | 0.4 |
| Asymptomatic | 77 (13%) | 3,272 (43%) | <0.001 |

\* 3 missing values not included
\*\* 107 missing values not included

We observed age variations in the symptomatology associated with being seropositive. In children <18 years old, anosmia and/or dysgeusia (OR 16.0, 95%CI 7.0-36.2), cough and/or fever (OR 1.8, 95%CI 1.1-3.1) and systemic symptoms (OR 1.9, 95%CI 1.0-3.2) were the groups of symptoms most strongly associated with seropositivity (Figure 1). On the contrary, among participants 65 years and older, dyspnea (OR 4.5, 95%CI 2.4-7.8), digestive symptoms (OR 3.1, 95%CI 1.7-5.2) and upper airways symptoms (OR 2.5, 95%CI 1.6-4.9) were more strongly associated with seropositivity than in the other age categories, although CIs overlap. In all age categories, those not reporting symptoms were less likely to be seropositive, although this association was strongest in the 18-64 years old age group (OR 0.17, 95% CI 0.12-0.23) and weakest in children (OR 0.55, 95% CI 0.30-0.97).

**Figure 1.**
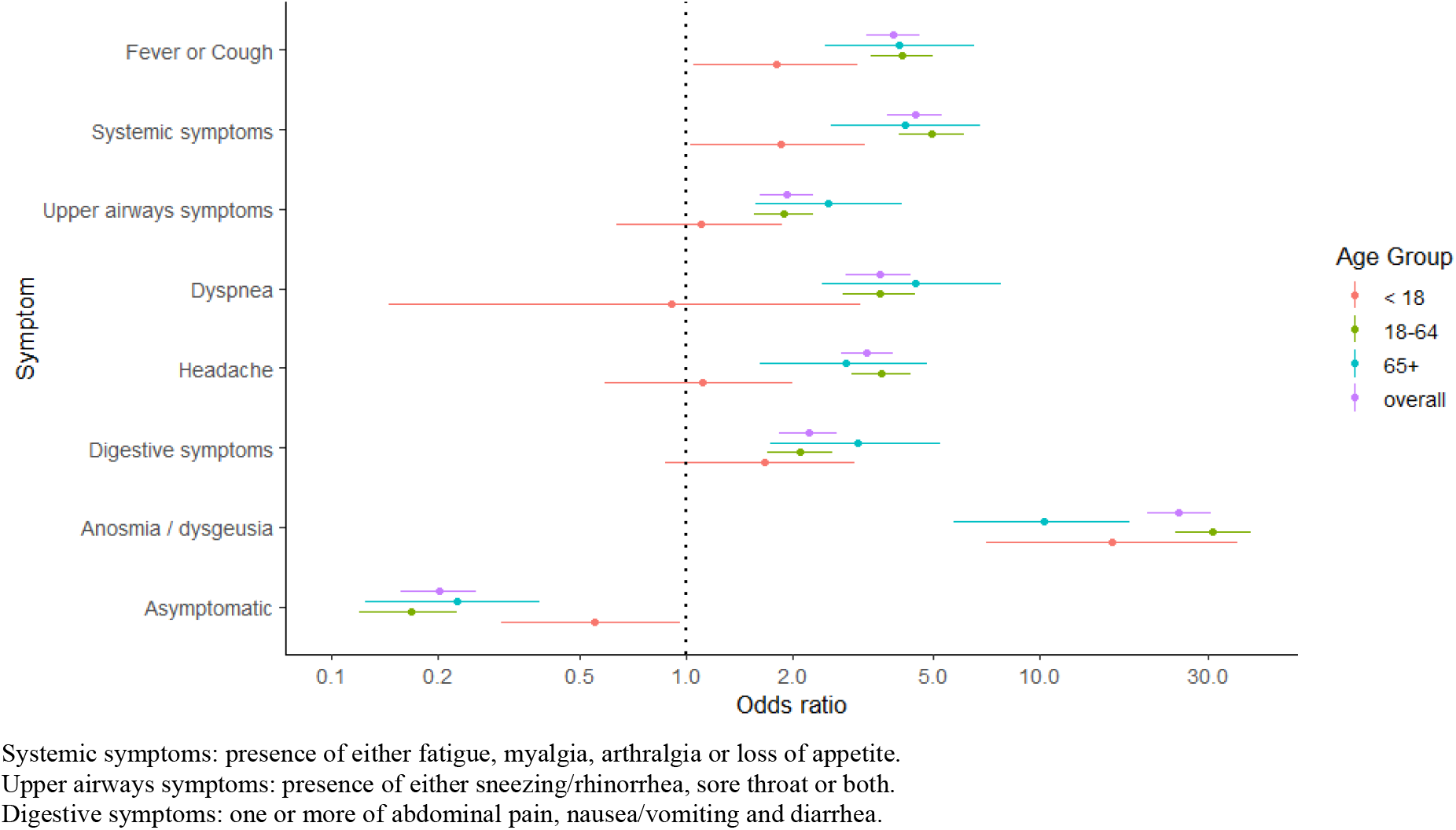
Age-stratified univariate odds ratios of seropositivity according to symptoms. Error bars correspond to 95% confidence intervals of odds ratios.

In the multivariate analysis with co-adjustment of symptoms we observed that nausea and/or vomiting, abdominal pain, and sore throat were negatively associated with being seropositive (Table S11).

Results for seroprevalence, risk factor and symptom analyses were mostly consistent when using the IFA confirmatory test (Supplement Tables S3, S10, S13 and S14). Frequency of symptoms and their association with being seropositive was also generally similar when considering only participants with one symptomatic episode at most (Supplement Tables S15 and S16).

## Discussion

This study including 8344 participants recruited between April and June 2020 confirms preliminary results showing that less than 10% of the general population of the canton of Geneva had developed anti-SARS-CoV-2 antibodies over the course of the first epidemic wave.(4) Consistent with our previously published interim analysis, children under 10 years old and individuals over 65 years old had the lowest risk of being seropositive, with males at higher risk than females. We did not identify clear associations between socioeconomic variables and serological status. Smoking appeared to reduce the odds of being SARS-CoV-2-seropositive, while no significant associations were found with other lifestyle and health-related risk factors. The reported symptom most strongly associated with seropositivity was anosmia/dysgeusia, followed by systemic symptoms such as loss of appetite, fever, fatigue and myalgia and/or arthralgia.

The average seroprevalence of 7.8% over our 12-week study period is in line with estimates from highly affected areas in other European countries.(2,5–7) The slight decrease in seroprevalence in the final weeks of our study may be due to sampling variation, caused by previously SARS-CoV-2-infected individuals being more prone to participate in the study during lockdown, which took place at an earlier phase of our study. Also, recruitment by postal mail of participants without a known email address (mostly elderly individuals) having begun during the second month of our study, older individuals came more frequently during the final month, thus possibly contributing to the decreasing seroprevalence. However, waning antibody levels after an initial infection might be another explanation for the observed decrease. Similar observations have been made in a large serosurvey in the United Kingdom (REACT2), where the adult population seroprevalence declined from 6.0% to 4.4% between June and September 2020.(3) Weekly fluctuations in the crude proportion of seropositive participants (Table S2) could not be attributed to any laboratory issues, demographic differences, or recruitment effects and may thus be attributed to random variations.

Consistent with previous serosurveys,(2) our study shows a significantly higher risk of infection in men (RR 1.22, 95% CrI: 1.06-1.40) than in women. This difference should be considered when evaluating the increased risk of complications and mortality in men found in previous research.(24) We also found the highest relative risk for infection in adults of working age, teenagers and preadolescents while children and older adults had lower relative risks and seroprevalence. While advanced age is now widely recognized as a risk factor for complications and increased mortality(25), the relationship between age and susceptibility to infection likely depends on multiple factors. Our study, like other serosurveys, was partly conducted during a lockdown. As such, it is likely that the measures in place affected adults, children and elderly individuals differently. Better social isolation and a strict application of preventive measures could have been more widely adopted by the elderly, out of fear of suffering from severe complications. Also, school closures may have contributed to the low seroprevalence observed in young children in our study, although recent household analyses based on this study suggest that children may be at reduced risk of infection even if exposed.(26) Adults of working age, on the other hand, could have had more difficulty effectively isolating from social contacts, since they were likely the ones to do grocery shopping and might have been essential workers, as evidenced by only a minority of participants (43.0%) having reported working remotely since the beginning of the pandemic. However, an immunological cause for the lower seroprevalence in young children and older individuals cannot be excluded at this stage.

Odds of seropositivity were significantly reduced for retired women and unemployed men compared to employed individuals of the same sex, after adjusting for age. Seroprevalence was also lower, although with borderline significance, for all retired and unemployed individuals. This could be due to these individuals having more flexibility in their daily routine allowing them to better self-isolate during the epidemic. While educational level was not significantly associated to the risk of infection, we did see trends of increasing association to seropositivity with increased educational levels, except for doctoral-level education. It is possible that the risk associated with each educational level is linked to the types of profession open to these individuals, which can be quite heterogenous, making it difficult to see significant differences. Finally, no significant association was found between median living-area-based income and SARS-CoV-2 seropositivity.

Similar to previous reports, we found that current smokers were less likely to be seropositive.(5) Several hypotheses exist, including the effects of smoking on the expression of the angiotensin-converting-enzyme 2 receptor in cells of the respiratory tract and the competition between nicotine and SARS-CoV-2 for the nicotinic acetylcholine receptor (nACfR), which acts as a co-receptor for viral cell entry.(27) Further work is needed to understand the behaviors that may contribute to this association, such as potential risk-averting behavior in smokers out of fear of respiratory complications of COVID-19. Confirming previous rt-PCR-based studies,(28) in our sample, anosmia was frequently reported in seropositive (44%) but rarely in seronegative participants (3%), with individuals reporting this symptom being almost 25 times more likely to be seropositive than those who did not. However, as previously reported (29,30), the correlation was weaker among individuals aged 65 years and older, possibly because the senses of taste and smell tend to decrease with age, thus making the symptom less discriminant.(31,32) A recent meta-analysis based on rt-PCR diagnosis estimated the proportion of asymptomatic people infected with SARS-CoV-2 at 20% (95% CI 17– 25%) with a prediction interval of 3%–67%.(33) However, due to the short window of relatively high sensitivity of rt-PCR, not all asymptomatic individuals are captured by molecular testing. Also, individuals considered asymptomatic at time of rt-PCR are potentially just presymptomatic.(34) Serological data mostly elude these difficulties if the delay between symptoms and testing is long enough. In our sample, 13% of IgG-positive and 43% of seronegative individuals reported being asymptomatic. As such, the number of true asymptomatic SARS-CoV-2 seropositive individuals in our sample is likely underestimated, due to the possibility to report symptoms related to multiple episodes.

Our study found some age-related differences in terms of clinical presentation. In the less than 18-year-olds, only fever, cough and systemic symptoms were associated with seropositivity, whereas in adults, all symptom groups were significantly associated with seropositivity. Also, while being asymptomatic reduces the odds of seropositivity almost five-fold in the overall study population, odds were only twice reduced in children, with a higher percentage of asymptomatic positive children than in positive adults (37% vs. 10%, table S11).

Our study comes with a number of limitations. The Bus Santé source population was originally composed of individuals between 20 and 75 years old, explaining the lower number of participants aged less than 18 and over 75 years in our final sample, despite the inclusion of primary participants’ household members. Delayed recruitment by ordinary mail of participants without an email address on file is likely to have had an influence on weekly seroprevalence results. Further, the association of income level with a seropositive status could not be assessed exactly, as we did not collect individual participants’ income, but estimated income based on the place of residency. Memory bias has likely influenced symptoms analysis,as participants were asked to report symptoms having occurred up to six months earlier. Second, symptoms reported by IgG-positive participants might have been caused by other pathogens as 1644 participants reported multiple episodes. Finally, the severity of the disease was not addressed in this study.

Our study determined that the average seroprevalence of SARS-CoV-2 in the canton of Geneva over the course of the first wave was 7.8 %, leaving most of the population naïve to the virus.(35) Our risk factor analysis indicates that while some socio-economic disparities in susceptibility to the infection do exist, they are likely more complex than what we previously thought. Older individuals seemed to be protected from infection during the first wave, perhaps due to targeted preventive measures. Some important observations could be made regarding differences in symptoms among age groups, such as the proportion of asymptomatic infections. However, future serosurveys should focus on larger samples of young children and the elderly to yield more detailed age-specific results regarding symptoms.

As sufficient vaccination of the population will likely take time, repeated seroprevalence surveys will be crucial to monitor the progression of the epidemic. Following up seropositive individuals will allow for determining the extent of the immune protection conferred by specific antibodies against re-infection by SARS-CoV-2, as well as provide insight on the prevalence of “long COVID”. Also, better characterization of the socio-demographic risk factors for infection is needed to further improve prevention strategies. These measures will be of paramount importance to guide public health policy in order to effectively manage future outbreaks of COVID-19.

## Data Availability

The authors confirm that the data supporting the findings of this study are available within the article and its supplementary materials.

## Acknowledgements

This study would not have been possible without the instrumental and passionate contribution of the staff of the Unit of Population Epidemiology of the HUG Primary Care Division, Geneva University Hospitals, Geneva, Switzerland (Natacha Noel, Caroline Pugin, Jane Portier, Barinjaka Rakotomiaramanana, Natalie Francioli, Paola d’Ippolito, Chantal Martinez, Francesco Pennacchio, Benjamin Emery, Zoé Waldman, Magdalena Schellongova, Prune Collombet, and Attilio Picazio), of the team of the Division of Laboratory Medicine, Geneva University Hospitals (Géraldine Poulain and Pierre Lescuyer), of the Geneva University Hospitals staff affected to our unit during the COVID-19 emergency (Lilas Salzmann-Bellard, Mélanie Seixas Miranda, Yasmina Malim, Acem Gonul, and Odile Desvachez), Loan Mattera and all the personnel of Campus Biotech, and finally without the invaluable work of the medical students who have invested their time and energy in this project (Jonathan Barbolini, Eugénie de Weck, Natacha Michel, Emmanuelle Mohbat, Irine Sakvarelidze, Céline Eelbode, Sultan Bahta, Céline Dubas, Lina Hassar, Melis Kir, Hugo-Ken Oulevey, Kourosh Massiha, Manon Will, Natacha Vincent, Fanny Lombard, Alioucha Davidovic, Benoit Favre, Amélie Mach, Eva Marchetti, Sophie Cattani, Joséphine Duc, Julie Guérin, Soraya Maret, Francesca Hovagemyan, Antoine Daeniker, and Rebecca Buetzberger). We also thank Markus Hoffmann and Stefan Pöhlmann (Georg-August-Universität Göttingen, Göttingen, Germany) for providing the SARS-CoV-2 spike expression vector used in the recombinant immunofluorescence, the Geneva University Hospitals archives, Human Neuroscience Platform, Foundation Campus Biotech Geneva for allowing us to use their premises for recruitment, and all the participants of the Bus Santé study and their household members. This study was funded by the Swiss Federal Office of Public Health, the Corona Immunitas research program of the Swiss School of Public Health, the Fondation de Bienfaisance du Groupe Pictet, the Fondation Ancrage, the Fondation Privée des HUG, and the Center for Emerging Viral Diseases. ASA is funded by the US National Institutes of Health (R01 AI135115) and The Bill & Melinda Gates Foundation (OPP1191944).

## Notes

### Competing Interest Statement

The authors have declared no competing interest.

### Author Declarations

The study protocol was approved by the Cantonal Research Ethics Commission of Geneva, Switzerland (CER16-363).

## References

1. Chen X, Chen Z, Azman AS, Deng X, Chen X, Lu W, et al. Serological evidence of human infection with SARS-CoV-2: a systematic review and meta-analysis. medRxiv. 2020 Oct 29;

2. Lai C-C, Wang J-H, Hsueh P-R. Population-based seroprevalence surveys of anti-SARS-CoV-2 antibody: An up-to-date review. International Journal of Infectious Diseases. 2020 Oct 9;0(0).

3. Ward H, Cooke G, Atchison C, Whitaker M, Elliott J, Moshe M, et al. Declining prevalence of antibody positivity to SARS-CoV-2: a community study of 365,000 adults. medRxiv. 2020 Oct 27;2020.10.26.20219725.

4. Stringhini S, Wisniak A, Piumatti G, Azman AS, Lauer SA, Baysson H, et al. Seroprevalence of anti-SARS-CoV-2 IgG antibodies in Geneva, Switzerland (SEROCoV-POP): a population-based study. The Lancet. 2020 Aug 1;396(10247):313–9.

5. Carrat F, Lamballerie X de, Rahib D, Blanche H, Lapidus N, Artaud F, et al. Seroprevalence of SARS-CoV-2 among adults in three regions of France following the lockdown and associated risk factors: a multicohort study. medRxiv. 2020 Sep 18;2020.09.16.20195693.

6. Vena A, Berruti M, Adessi A, Blumetti P, Brignole M, Colognato R, et al. Prevalence of Antibodies to SARS-CoV-2 in Italian Adults and Associated Risk Factors. Journal of Clinical Medicine. 2020 Sep;9(9):2780.

7. Pollán M, Pérez-Gómez B, Pastor-Barriuso R, Oteo J,Hernán MA, Pérez-Olmeda M, et al. Prevalence of SARS-CoV-2 in Spain (ENE-COVID): a nationwide, population-based seroepidemiological study. The Lancet. 2020 Aug 22;396(10250):535–44.

8. Vu SL, Jones G, Anna F, Rose T, Richard J-B, Bernard-Stoecklin S, et al. Prevalence of SARS-CoV-2 antibodies in France: results from nationwide serological surveillance. medRxiv. 2020 Oct 21;2020.10.20.20213116.

9. Rozenfeld Y, Beam J, Maier H, Haggerson W, Boudreau K, Carlson J, et al. A model of disparities: risk factors associated with COVID-19 infection. International Journal for Equity in Health. 2020 Jul 29;19(1):126.

10. de Lusignan S, Dorward J, Correa A, Jones N, Akinyemi O, Amirthalingam G, et al. Risk factors for SARS-CoV-2 among patients in the Oxford Royal College of General Practitioners Research and Surveillance Centre primary care network: a cross-sectional study. Lancet Infect Dis. 2020;20(9):1034–42.

11. Martins LD, da Silva I, Batista WV, de Fátima Andrade M, Dias de Freitas E, Martins JA. How socio-economic and atmospheric variables impact COVID-19 and Influenza outbreaks in tropical and subtropical regions of Brazil. Environ Res. 2020 Sep 15;110184.

12. Raifman MA, Raifman JR. Disparities in the Population at Risk of Severe Illness From COVID- 19 by Race/Ethnicity and Income. Am J Prev Med. 2020 Jul;59(1):137–9.

13. Raisi-Estabragh Z, McCracken C, Bethell MS, Cooper J, Cooper C, Caulfield MJ, et al. Greater risk of severe COVID-19 in Black, Asian and Minority Ethnic populations is not explained by cardiometabolic, socioeconomic or behavioural factors, or by 25(OH)-vitamin D status: study of 1326 cases from the UK Biobank. J Public Health (Oxf). 2020 Aug 18;42(3):451–60.

14. Ward H, Atchison CJ, Whitaker M, Ainslie KEC, Elliott J, Okell LC, et al. Antibody prevalence for SARS-CoV-2 in England following first peak of the pandemic: REACT2 study in 100,000 adults. medRxiv. 2020 Aug 21;2020.08.12.20173690.

15. Menachemi N, Yiannoutsos CT, Dixon BE, Duszynski TJ, Fadel WF, Wools-Kaloustian KK, et al. Population Point Prevalence of SARS-CoV-2 Infection Based on a Statewide Random Sample — Indiana, April 25–29, 2020. MMWR Morb Mortal Wkly Rep. 2020 Jul 24;69(29):960–4.

16. Xu X, Sun J, Nie S, Li H, Kong Y, Liang M, et al. Seroprevalence of immunoglobulin M and G antibodies against SARS-CoV-2 in China. Nature Medicine. 2020 Aug;26(8):1193–5.

17. COVID-19 - Situation Suisse : Évolution dans le temps [Internet]. [cited 2020 Sep 20]. Available from: https://covid-19-schweiz.bagapps.ch/fr-2.html

18. Nouveau coronavirus : situation en Suisse [Internet]. Office fédéral de la santé publique OFSP. [cited 2020 May 22]. Available from: https://www.bag.admin.ch/bag/fr/home/krankheiten/ausbrueche-epidemien-pandemien/aktuelle-ausbrueche-epidemien/novel-cov/situation-schweiz-und-international.html

19. Guessous I, Gaspoz JM, Theler JM, Wolff H. High prevalence of forgoing healthcare for economic reasons in Switzerland: a population-based study in a region with universal health insurance coverage. Prev Med. 2012 Nov;55(5):521–7.

20. RS 818.101.24 Ordonnance 2 du 13 mars 2020 sur les mesures destinées à lutter contre le coronavirus (COVID-19) (Ordonnance 2 COVID-19) [Internet]. [cited 2020 Nov 18]. Available from: https://www.admin.ch/opc/fr/classified-compilation/20200744/index.html

21. Meyer B, Torriani G, Yerly S, Mazza L, Calame A, Arm-Vernez I, et al. Validation of a commercially available SARS-CoV-2 serological immunoassay. Clinical Microbiology and Infection. 2020 Oct 1;26(10):1386–94.

22. Statistiques cantonales - République et canton de Genève [Internet]. [cited 2020 Oct 1]. Available from: https://www.ge.ch/statistique/

23. R Foundation for Statistical Computing, Vienna, Austria. R: A language and environment for statistical computing. [Internet]. [cited 2020 Oct 1]. Available from: https://www.r-project.org/

24. Kopel J, Perisetti A, Roghani A, Aziz M, Gajendran M, Goyal H. Racial and Gender-Based Differences in COVID-19. Front Public Health. 2020 Jul 28;8.

25. The Novel Coronavirus Pneumonia Emergency Response Epidemiology Team. The epidemiological characteristics of an outbreak of 2019 novel coronavirus diseases (COVID-19) — China, 2020. Vital Sureillance. 2020 Feb 10;41(2):145–51.

26. Bi Q, Lessler J, Eckerle I, Lauer SA, Kaiser L, Vuilleumier N, et al. Household Transmission of SARS-COV-2: Insights from a Population-based Serological Survey. medRxiv. 2020 Nov 4;2020.11.04.20225573.

27. Grundy EJ, Suddek T, Filippidis FT, Majeed A, Coronini-Cronberg S. Smoking, SARS-CoV-2 and COVID-19: A review of reviews considering implications for public health policy and practice. Tob Induc Dis. 2020;18:58.

28. Haehner A, Draf J, Dräger S, de With K, Hummel T. Predictive Value of Sudden Olfactory Loss in the Diagnosis of COVID-19. ORL J Otorhinolaryngol Relat Spec. 2020;82(4):175–80.

29. von Bartheld CS, Hagen MM, Butowt R. Prevalence of Chemosensory Dysfunction in COVID- 19 Patients: A Systematic Review and Meta-analysis Reveals Significant Ethnic Differences. ACS Chem Neurosci. 2020 07;11(19):2944–61.

30. Agyeman AA, Chin KL, Landersdorfer CB, Liew D, Ofori-Asenso R. Smell and Taste Dysfunction in Patients With COVID-19: A Systematic Review and Meta-analysis. Mayo Clin Proc. 2020;95(8):1621–31.

31. Methven L, Allen VJ, Withers CA, Gosney MA. Ageing and taste. Proc Nutr Soc. 2012 Nov;71(4):556–65.

32. Doty RL, Kamath V. The influences of age on olfaction: a review. Front Psychol. 2014 Feb 7;5.

33. Buitrago-Garcia D, Egli-Gany D, Counotte MJ, Hossmann S, Imeri H, Ipekci AM, et al. Occurrence and transmission potential of asymptomatic and presymptomatic SARS-CoV-2 infections: A living systematic review and meta-analysis. PLOS Medicine. 2020 Sep 22;17(9):e1003346.

34. Lee S, Meyler P, Mozel M, Tauh T, Merchant R. Asymptomatic carriage and transmission of SARS-CoV-2: What do we know? Can J Anaesth. 2020 Jun 2;1–7.

35. COVID-19 - bilan épidémiologique hebdomadaire [Internet]. ge.ch. [cited 2020 Dec 5]. Available from: https://www.ge.ch/node/19696

