## Supplemental materials for "Seroprevalence of anti-SARS-CoV-2 IgG antibodies, risk factors for infection and associated symptoms in Geneva, Switzerland: a population-based study"

**Supplementary Materials**

Figure S1. Flow chart for the inclusion of participants in the SEROCOV-POP study

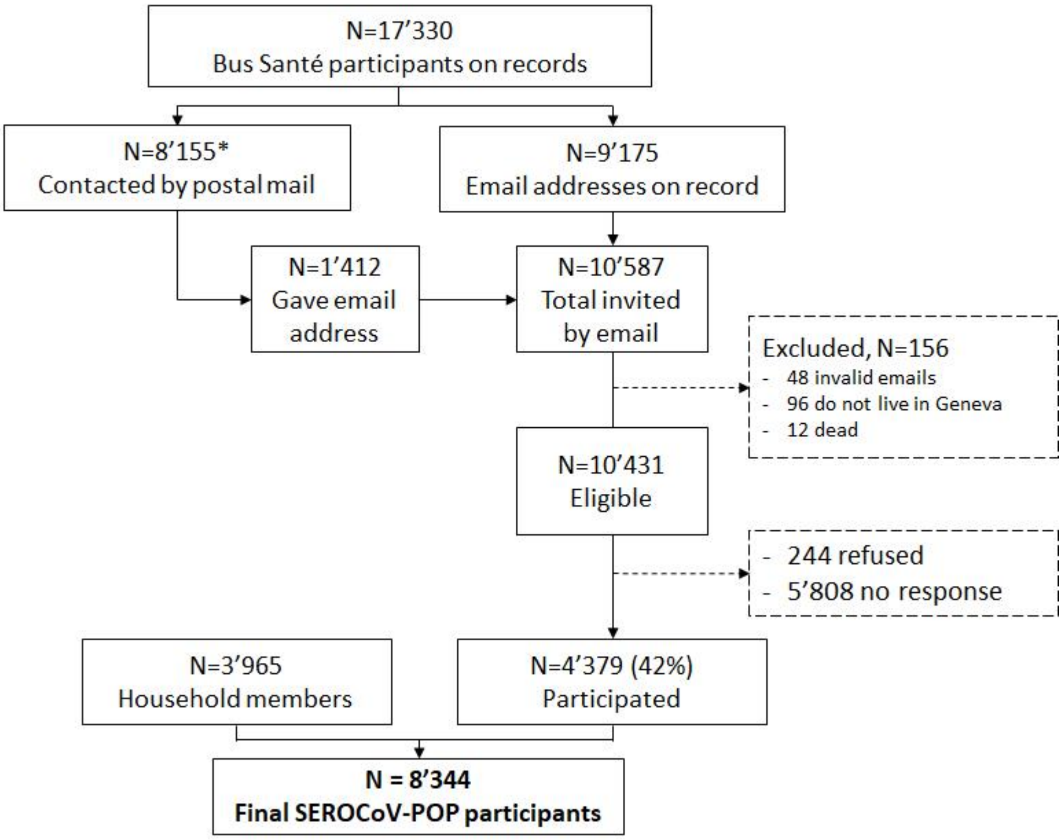


*: mostly non-reachable as records could go as far as 30 years back

Table S1. Sex, age and level of education distribution in the SEROCoV-POP sample and the population of Geneva

| | SEROCoV-POP sample | Geneva population |
|---|---|---|
| **Sex** | | |
| Female | 53.5% | 51.5% |
| Male | 46.5% | 48.5% |
| **Age category** | | |
| 5-9 | 3.3% | 5.2% |
| 10-19 | 9.6% | 10.5% |
| 20-49 | 35.2% | 43.3% |
| 50-64 | 32.3% | 19.4% |
| 65-74 | 14.3% | 8.0% |
| 75-89 | 5.2% | 7.5% |
| 90+ | 0.1% | 1.1% |
| **Education level** | | |
| Mandatory | 6.8% | 28.1% |
| Secondary | 35.6% | 32.5% |
| Tertiary | 57.7% | 39.3% |

### Section 1: seroprevalence and relative risk by age and sex

We follow the Bayesian regression methodology developed in Stringhini et al, 2020. We here account for age, sex, houshold-level effects, and the sensitivity and specificity of the ELISA assay in the seropositive probability of each serosurvey participant. Following the same notation as in Stringhini et al, we have that:

$$x_i \sim Bernoulli(p_i\theta^+ + (1-p_i)(1-\theta^-)),$$
$$logit(p_i) = \alpha_0 + \alpha_h + \beta_{age}\, age_i + \beta_{sex}\, sex_i,$$
$$\alpha_h \sim Normal(0, \sigma^2),$$
$$x^+ \sim Binomial(n^+, \theta^+),$$
$$x^- \sim Binomial(n^-, \theta^-),$$

where $x_i$ is the result of the IgG ELISA (in the main text), $p_i$ is the probability that participant $i$ is seropositive, $\theta^+$and $\theta^-$ the sensitivity and specificity of the ELISA assay. The later are determined with the set of $n^+$ RT-PCR positive controls from the lab validation study, of which $x^+$ tested positive, and $n^-$ pre-pandemic negative controls, of which $x^-$ tested positive. The logistic seropositivity probability model accounts for a household-level random effect h with variance 2, the age class and the sex of participants through coefficients $\beta_{age}$, $\beta_{sex}$ and an intercept $\alpha_0$. As in Stringhini et al. 2020, We used weak Normal (0,1) priors for the logistic regression coefficients β. The prior on the standard deviation of the household effect,σ, was flat from 0 to infinity. We implemented this model in the Stan probabilistic programming language and used therstanpackagein R to run the model and analyse outputs. We ran 5,000 iterations (4 chains with 1,500 iterations each with250 for warm-up) and assessed convergence visually and using the R-hat statistic.

Following Stringhini et al. 2020, we calculated the relative risk (RR) of being seropositive for each subset using the posterior draws for each logistic regression coefficient and integrating across the household random effect. We selected age 18–49 years as the reference age group because it had the largest sample size.

Sensitivity analysis covers the use of the IFA result to define seropositivity. Seroprevalence estimates for alternative age categories are also provided.

Table S2 Crude percentage of seropositive participants: (a) per week and (b) per month, between April and June 2020.

(a)

| Week | Percent positive (N/Ntot) | 95% CI* |
|---|---|---|
| 1 | 3.5 (12/339) | 2.0 - 6.1 |
| 2 | 6.0 (28/468) | 4.1 - 8.5 |
| 3 | 10.4 (61/589) | 8.1 - 13.1 |
| 4 | 5.8 (35/605) | 4.2 - 8.0 |
| 5 | 10.6 (82/774) | 8.6 - 13.0 |
| 6 | 5.9 (45/759) | 4.4 - 7.9 |
| 7 | 10.1 (74/733) | 8.1 - 12.5 |
| 8 | 5.2 (35/668) | 3.8 - 7.2 |
| 9 | 7.2 (47/657) | 5.4 - 9.4 |
| 10 | 5.0 (42/846) | 3.7 - 6.7 |
| 11 | 7.6 (65/858) | 6.0 - 9.6 |
| 12 | 5.9 (62/1046) | 4.6 - 7.5 |

*95% confidence intervals using the Agresti-Coull method

(b)

| Month | Percent positive (N/Ntot) | 95% CI* |
|---|---|---|
| April | 6.7 (130/1939) | 5.7 - 7.9 |
| May | 8.1 (242/2996) | 7.2 - 9.1 |
| June | 6.3 (216/3407) | 5.6 - 7.2 |

*95% confidence intervals using the Agresti-Coull method

Table S3. Seroprevalence and seropositivity relative risk estimates with for 10-year age classes

| | SARS-CoV-2 serology test result | | Seroprevalence (95% CI) | Relative risk (95% CI) | p-value |
|---|---|---|---|---|---|
| | Positive | Negative | | | |
| **Age group, years** | | | | | |
| 4-9 (n=274) | 9 (3.3%) | 265 (96.7%) | 4.1 (2.0 - 6.7) | **0.48 (0.24 - 0.80)** | **0.0044** |
| 10-19 (n=802) | 65 (8.1%) | 737 (91.9%) | 7.9 (6.0 - 10.0) | **0.95 (0.71 - 1.2)** | 0.68 |
| 20-29 (n=811) | 88 (10.9%) | 723 (89.2%) | 10.9 (8.5 - 13.4) | **1.3 (1.0 - 1.7)** | **0.026** |
| 30-39 (n=782) | 67 (8.6%) | 715 (91.4%) | 9.3 (7.0 - 11.8) | **1.1 (0.84 - 1.5)** | 0.42 |
| 40-49 (n=1341) | 108 (8.1%) | 1233 (92.0%) | 8.7 (7.0 - 10.6) | **1.1 (0.8 - 1.3)** | 0.67 |
| 50-59 (n=1944) | 138 (7.1%) | 1806 (92.9%) | 8.3 (6.9 - 9.9) | **--** | -- |
| 60-69 (n=1374) | 68 (5.0%) | 1306 (95.1%) | 5.6 (4.1 - 7.1) | **0.67 (0.50 - 0.88)** | **0.0036** |
| >70 (n=1016) | 47 (4.6%) | 969 (95.4%) | 5.2 (3.7 - 6.9) | **0.64 (0.44 - 0.87)** | **0.0032** |
| **Sex** | | | | | |
| Female (n=4465) | 305 (7.9%) | 3574 (92.1%) | 8.8 (7.6 - 10.1) | **1.2 (1.1-1.4)** | **0.0032** |
| Male (n=3879) | 285 (6.4%) | 4180 (93.6%) | 7.1 (6.1 - 8.2) | -- | -- |
| **Overall** (n=8344) | 590 (7.1%) | 7754 (92.9%) | 7.9 (6.9 – 9.0) | -- | -- |

Table S4. Seroprevalence and seropositivity relative risk estimates with IFA

| | SARS-CoV-2 serology test result | | Seroprevalence (95% CI) | Relative risk (95% CI) | p-value |
|---|---|---|---|---|---|
| | Positive | Negative | | | |
| **Age group, years** | | | | | |
| 4-9 (n=274) | 6 (2.2%) | 268 (97.8%) | 3.2 (1.5 - 5.4) | **0.39 (0.19 - 0.65)** | **0** |
| 10-17 (n=628) | 44 (7.0%) | 584 (93.0%) | 6.3 (4.7 - 8.3) | **0.77 (0.57 - 1.0)** | 0.057 |
| 18-49 (n=3108) | 253 (8.1%) | 2855 (91.9%) | 8.2 (7.1 - 9.5) | **--** | -- |
| 50-64 (n=2694) | 168 (6.2%) | 2526 (93.8%) | 6.7 (5.6 - 7.9) | **0.82 (0.68 - 0.97)** | **0.022** |
| 65-74 (n=1196) | 44 (3.7%) | 1152 (96.3%) | 4.3 (3.1 - 5.7) | **0.53 (0.38 - 0.70)** | **0** |
| >75 (n=444) | 16 (3.6%) | 428 (96.4%) | 4.7 (2.8 - 7.1) | **0.57 (0.34 - 0.87)** | **0.0084** |
| **Sex** | | | | | |
| Female (n=4465) | 256 (6.6%) | 3623 (93.4%) | 7.0 (6.0 - 8.0) | 1.1 (0.91 - 1.2) | 0.52 |
| Male (n=3879) | 275 (6.2%) | 4190 (93.8%) | 6.6 (5.7 - 7.6) | -- | -- |
| **Overall** (n=8344) | 531 (6.4%) | 7813 (93.6%) | 6.8 (6.0 - 7.7) | -- | -- |

Table S5. Seroprevalence and seropositivity relative risk estimates without first three weeks

| | SARS-CoV-2 serology test result | | Seroprevalence (95% CI) | Relative risk (95% CI) | p-value |
|---|---|---|---|---|---|
| | Positive | Negative | | | |
| **Age group, years** | | | | | |
| 4-9 (n=215) | 8 (3.7%) | 207 (96.3%) | 5.1 (2.5 - 8.5) | **0.54 (0.27 - 0.90)** | **0.0132** |
| 10-17 (n=498) | 38 (7.6%) | 460 (92.4%) | 7.8 (5.5 - 10.4) | **0.83 (0.59 - 1.1)** | 0.2116 |
| 18-49 (n=2494) | 221 (8.9%) | 2273 (91.1%) | 9.4 (7.9 - 10.9) | **--** | -- |
| 50-64 (n=2274) | 155 (6.8%) | 2119 (93.2%) | 7.51 (6.1 - 8.9) | **0.80 (0.65 - 0.97)** | **0.0216** |
| 65-74 (n=1054) | 50 (4.7%) | 1004 (95.3%) | 5.2 (3.7 - 6.9) | **0.56 (0.39 - 0.76)** | **0** |
| >75 (n=413) | 17 (4.1%) | 396 (95.9%) | 4.95 (2.9 - 7.5) | **0.54 (0.31 - 0.82)** | **0.0036** |
| **Sex** | | | | | |
| Female (n=3719) | 239 (6.4%) | 3480 (93.6%) | 7.2 (6.0 - 8.4) | -- | |
| Male (n=3229) | 250 (7.7%) | 2979 (92.3%) | 8.6 (7.1 – 10.0) | 1.2 (1.0 - 1.4) | 0.0568 |
| **Overall** (n=6948) | 489 (7.0%) | 6459 (93.0%) | 7.8 (6.7 - 8.9) | -- | -- |

To account for potentially increasing seroprevalence during the first three study weeks due to ongoing transmission, as shown in our previous analysis,(4) we refit the models excluding the first three weeks. However, the results show a stable overall seroprevalence estimate (7.8%, 95% CrI 6.7-8.9) compared to the main analysis including all 12 weeks (7.8 %, 95% CrI 6.8-8.9).

Table S6. Seroprevalence and seropositivity relative risk estimates by school grade

| | SARS-CoV-2 serology test result | | Seroprevalence (95% CI) | Relative risk (95% CI) | p-value |
|---|---|---|---|---|---|
| | Positive | Negative | | | |
| **Age group, years** | | | | | |
| 4-11 (n=404) | 20 (5.0%) | 384 (95.1%) | 5.6 (3.5 - 8.2) | **0.60 (0.37 - 0.88)** | **0.0084** |
| 12-14 (n=229) | 17 (7.4%) | 212 (92.6%) | 7.6 (4.5 - 11.2) | 0.81 (0.49 - 1.2) | 0.31 |
| 15-19 (n=443) | 37 (8.4%) | 406 (91.7%) | 8-0 (5.6 - 10.8) | 0.86 (0.59 - 1.2) | 0.36 |
| 20-25 (n=541) | 60 (11.1%) | 481 (88.9%) | 10.8 (8.2 - 13.7) | 1.2 (0.87 - 1.5) | 0.27 |
| 26-49 (n=2393) | 203 (8.5%) | 2190 (91.5%) | 9.3 (7.9 - 10.8) | -- | -- |
| 50-64 (n=2694) | 180 (6.7%) | 2514 (93.3%) | 7.5 (6.3 - 8.9) | **0.81 (0.66 - 0.98)** | **0.034** |
| 65-74 (n=1196) | 56 (4.7%) | 1140 (95.3%) | 5.4 (4.0 – 7.0) | **0.58 (0.42 - 0.77)** | **0.0004** |
| >75 (n=444) | 17 (3.8%) | 427 (96.2%) | 4.6 (2.7 - 7.0) | **0.51 (0.29 - 0.78)** | **0.0012** |
| **Sex** | | | | | |
| Female (n=4465) | 285 (6.4%) | 4180 (93.6%) | 7.1 (6.0 - 8.1) | -- | -- |
| Male (n=3879) | 305 (7.9%) | 3574 (92.1%) | 8.8 (7.6 - 10.1) | **1.2 (1.1 - 1.4)** | **0.008** |
| **Overall** (n=8344) | 590 (7.1%) | 7754 (92.9%) | 7.9 (6.9 - 8.9) | -- | -- |

**Section 2: Socio-economic factors**

Figure S2. Frequency plot of missingness and most-reported combinations of missingness.

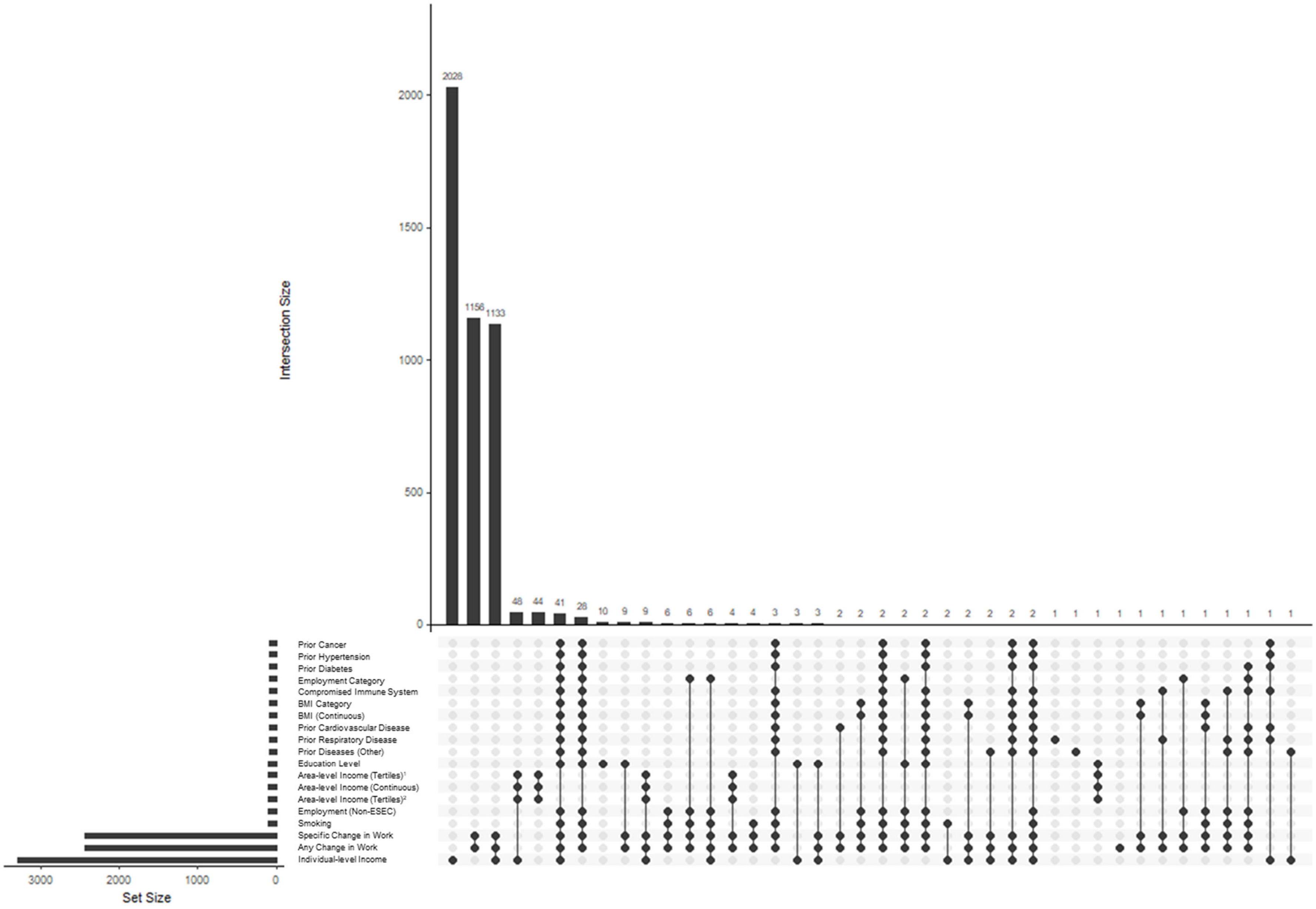


[1]Tertile categories for area-level income with tertile 1 as reference.
[2]Tertile categories for area-level income with tertile 3 as reference.

Figure S3. Facet plot of missingness based on negative/positive IgG test for SARS-CoV-2 infection among all variables included in multiple imputation.

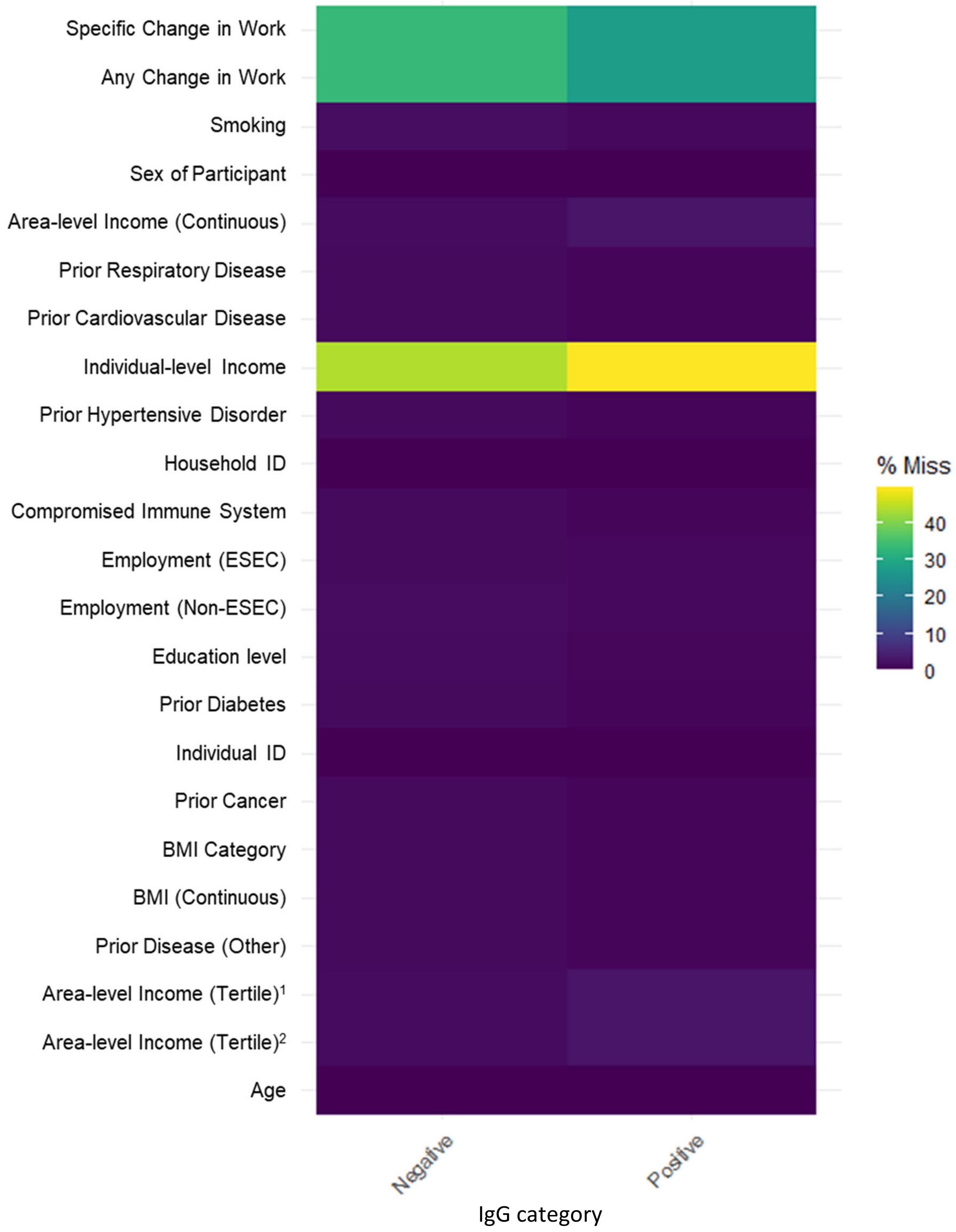


[1]Tertile categories for area-level income with tertile 1 as reference.
[2]Tertile categories for area-level income with tertile 3 as reference.

Table S7. Distribution of sociodemographic variables following multiple imputation.

| | Overall | SARS-CoV-2 serology test result | | Adjusted probabilites* | p-value* |
|---|---|---|---|---|---|
| | | Negative | Positive | | |
| **Employment Status** | | | | | |
| Employed | 3680 (49.4%) | 3392 (92.2%) | 288 (7.8%) | 7.3% | *Ref* |
| Unemployed | 566 (7.6%) | 532 (94.0%) | 34 (6.0%) | 5.8% | 0.68 |
| Student | 693 (9.3%) | 623 (89.9%) | 70 (10.1%) | 7.0% | 1.00 |
| Retired | 1676 (22.5%) | 1604 (95.7%) | 72 (4.3%) | 5.6% | 0.45 |
| Freelance/Other | 827 (11.1%) | 760 (91.9%) | 67 (8.1%) | 8.3% | 0.87 |
| **Occupational Position** | | | | | |
| Professionals/Managers | 760 (10.2%) | 697 (91.7%) | 63 (8.3%) | 7.9% | *Ref* |
| Higher grade WCW | 1163 (15.6%) | 1075 (92.4%) | 88 (7.6%) | 7.2% | 1.00 |
| Independents | 169 (2.3%) | 158 (93.5%) | 11 (6.5%) | 6.4% | 1.00 |
| Lower grade WCW | 938 (12.6%) | 863 (92.0%) | 75 (8.0%) | 7.8% | 1.00 |
| BCW | 376 (5.1%) | 349 (92.8%) | 27 (7.2%) | 6.3% | 0.98 |
| Other or N/A | 1101 (14.8%) | 1010 (91.7%) | 91 (8.3%) | 7.9% | 1.00 |
| **Change in work** | | | | | |
| None | 1604 (32.1%) | 1482 (92.4%) | 122 (7.6%) | 7.7% | *Ref* |
| Any | 3397 (67.9%) | 3133 (92.2%) | 264 (7.8%) | 7.6% | 0.88 |
| **Specific Change in Work** | | | | | |
| Stopped activities | 866 (11.6%) | 804 (92.8%) | 62 (7.2%) | 7.0% | 1.00 |
| Telework | 2983 (40.0%) | 2777 (93.1%) | 206 (6.9%) | 6.9% | 1.00 |
| Took sick leave | 73 (1.0%) | 54 (74.0%) | 19 (26.0%) | 24.1% | <0.0001 |
| Unemployed | 258 (3.5%) | 245 (95.0%) | 13 (5.0%) | 5.6% | 0.97 |
| Other | 650 (8.7%) | 600 (92.3%) | 50 (7.7%) | 6.6% | 1.00 |
| **Area-based income – Single (CHF)** | | | | | |
| >37000 | 396 (5.3%) | 367 (92.7%) | 29 (7.3%) | 7.2% | *Ref* |
| >=37000& <68000 | 6799 (91.4%) | 6316 (92.9%) | 483 (7.1%) | 6.7% | 0.97 |
| >=68000 | 247 (3.3%) | 228 (92.3%) | 19 (7.7%) | 7.5% | 0.99 |
| **Educational Status** | | | | | |
| Mandatory education only | 481 (6.5%) | 453 (94.2%) | 28 (5.8%) | 5.2% | *Ref* |
| Apprenticeship | 1132 (15.2%) | 1070 (94.5%) | 62 (5/5%) | 5.8% | 1.00 |
| Secondary education | 1376 (18.5%) | 1273 (92.5%) | 103 (7.5%) | 6.9% | 0.79 |
| Haute Ecole/ University | 3686 (49.5%) | 3395 (92.1%) | 291 (7.9%) | 7.7% | 0.35 |
| Doctoral education | 412 (5.5%) | 389 (94.4%) | 23 (5.6%) | 5.4% | 1.00 |
| Other | 355 (4.8%) | 311 (93.2%) | 24 (6.8%) | 7.1% | 0.87 |
| **Smoking** | | | | | |
| Never | 5010 (67.3%) | 4611 (92.0%) | 399 (8.0%) | 7.7% | *Ref* |
| Former | 1296 (17.4%) | 1213 (93.6%) | 83 (6.4%) | 6.9% | 0.60 |
| Current | 1136 (15.3%) | 1087 (95.7%) | 49 (4.3%) | 3.7% | <0.0001 |
| **BMI Category** | | | | | |
| Underweight | 395 (5.3%) | 369 (93.4%) | 26 (6.6%) | 5.9% | 0.85 |
| Normal weight | 4289 (57.6%) | 3974 (92.7%) | 315 (7.3%) | 7.0% | *Ref* |
| Overweight | 2082 (28.0%) | 1941 (93.2%) | 141 (6.8%) | 6.8% | 1.00 |
| Obese | 676 (9.1%) | 627 (92.8%) | 49 (7.2%) | 7.6% | 0.93 |
| **Chronic Health Conditions** | | | | | |
| None | 5699 (76.6%) | 5269 (92.5%) | 430 (7.5%) | 7.0% | *Ref* |
| 1 chronic condition | 1210 (16.3%) | 1132 (93.6%) | 78 (6.4%) | 7.3% | 0.94 |
| 2+ chronic conditions | 439 (5.9%) | 419 (95.4%) | 20 (4.6%) | 5.6% | 0.61 |

*Ref* : Reference.
*Adjusted for age and sex using logistic regression.

Table S8. Socioeconomic associations with positive SARS-CoV-2 IgG test with multilevel multiple imputation by chained equation.

| | Overall sample[1] | | Women[2] | | Men[2] | |
|---|---|---|---|---|---|---|
| | OR (95% CI) | p-value | OR (95% CI) | p-value | OR (95% CI) | p-value |
| **Employment** | | | | | | |
| Employed | [Reference] | | [Reference] | | [Reference] | |
| Unemployed | 0.72 (0.46 – 1.1) | 0.13 | 0.91 (0.58 – 1.5) | 0.70 | 0.50 (0.23 – 1.1) | 0.73 |
| Student | 0.86 (0.57 – 1.3) | 0.47 | 1.17 (0.72 – 1.9) | 0.53 | 0.71 (0.41 – 1.2) | 0.20 |
| Retired | 0.73 (0.49 – 1.1) | 0.11 | **0.55 (0.33 – 0.94)** | **0.03** | 0.94 (0.59, 1.5) | 0.80 |
| Unemployed | 1.13 (0.81 – 1.6) | 0.47 | 1.35 (0.88, 2.1) | 0.17 | 0.98 (0.66, 1.5) | 0.94 |
| **Occupational position** | | | | | | |
| Professional/Manager | [Reference] | | [Reference] | | [Reference] | |
| Higher grade WCW | 0.87 (0.58, 1.3) | 0.50 | 1.50 (0.81, 2.8) | 0.20 | 0.65 (0.41, 1.0) | 0.08 |
| Independent | 0.77 (0.36, 1.7) | 0.51 | 1.28 (0.40, 4.2) | 0.68 | 0.65 (0.28, 1.5) | 0.32 |
| Lower grade WCW | 1.01 (0.67, 1.5) | 0.97 | 1.47 (0.79, 2.7) | 0.22 | 0.65 (0.28, 1.5) | 0.32 |
| BCW | 0.72 (0.41, 1.3) | 0.26 | 0.90 (0.33, 2.4) | 0.84 | 0.87 (0.53, 1.5) | 0.59 |
| Other or N/A | 1.03 (0.68, 1.5) | 0.90 | 1.63 (0.87, 3.0) | 0.13 | 0.73 (0.41, 1.3) | 0.27 |
| **Educational level** | | | | | | |
| Doctoral Education | [Reference] | | [Reference] | | [Reference] | |
| Haute Ecole/ University | 1.39 (0.82, 2.3) | 0.22 | 2.29 (0.91, 5.8) | 0.08 | 1.16 (0.66, 2.0) | 0.60 |
| Secondary education | 1.19 (0.68, 2.1) | 0.55 | 1.98 (0.76, 5.1) | 0.16 | 1.05 (0.56, 2.0) | 0.87 |
| Apprenticeship | 1.07 (0.60,1.9) | 0.82 | 1.71 (0.63, 4.6) | 0.29 | 1.05 (0.46, 1.6) | 0.64 |
| Mandatory education only | 0.94 (0.48, 1.8) | 0.86 | 1.41 (0.47, 4.2) | 0.54 | 0.93 (0.43, 2.0) | 0.85 |
| Other | 1.18 (0.59, 2.4) | 0.64 | 2.38 (0.82, 7.0) | 0.11 | 0.83 (0.35, 2.0) | 0.67 |
| **Area level income** | | | | | | |
| <37000 | [Reference] | | [Reference] | | [Reference] | |
| >=37000 and <68000 | 0.86 (0.44 – 1.7) | 0.65 | 1.61 (0.63 – 4.2) | 0.32 | 0.57 (0.25 – 1.3) | 0.18 |
| >=68000 | 0.99 (0.36 – 2.8) | 0.98 | 1.73 (0.44 – 6.8) | 0.43 | 0.69 (0.19, 2.5) | 0.57 |
| **Individual Income** | | | | | | |
| <2999 CHF/month | [Reference] | | [Reference] | | [Reference] | |
| 3000 to 4999 CHF/month | 2.62 (0.88 – 7.8) | 0.08 | 2.08 (0.59, 7.4) | 0.26 | 3.63 (0.42, 31.00) | 0.24 |
| 5000 to 6999 CHF/month | 2.76 (0.96, 7.9) | 0.06 | 2.20 (0.64, 7.5) | 0.21 | 4.01 (0.51, 31.9) | 0.19 |
| 7000 to 9499 CHF/month | 1.74 (0.61, 5.0) | 0.30 | 1.06 (0.30, 3.8) | 0.93 | 3.77 (0.49, 29.0) | 0.20 |
| 9500 to 13000 CHF/month | 2.04 (0.72, 5.8) | 0.18 | 8.49 (0.23, 3.1) | 0.80 | 5.31 (0.71, 39.8) | 0.10 |
| >13000 CHF/month | 3.14 (1.13, 8.7) | 0.03 | 2.13 (0.63, 7.2) | 0.22 | 5.92 (0.80, 43.9) | 0.08 |
| Don't know | 2.51 (0.88, 7.1) | 0.08 | 2.24 (0.66, 7.6) | 0.20 | 3.38 (0.44, 26.1) | 0.24 |
| **Smoking** | | | | | | N= |
| Never | Reference | | Reference | | Reference | |
| Former | 0.87 (0.65, 1.2) | 0.36 | 0.75 (0.50, 1.1) | 0.15 | 0.99 (0.70, 1.4) | 0.94 |
| Current | **0.48 (0.34, 0.7)** | **0.00004** | **0.45 (0.28, 0.7)** | **0.0001** | **0.51 (0.33, 0.8)** | **0.002** |
| **BMI** | | | | | | N= |
| Reference | Reference | | Reference | | Reference | |
| Underweight | 0.69 (0.42, 1.2) | 0.13 | 0.95 (0.59, 1.5) | 0.82 | 0.28 (0.07, 1.2) | 0.09 |
| Overweight | 1.00 (0.78, 1.3) | 0.99 | 0.90 (0.62, 1.3) | 0.55 | 1.01 (0.76, 1.4) | 0.93 |
| Obese | 1.22 (0.84, 1.8) | 0.29 | 1.52 (0.97, 2.4) | 0.07 | 0.83 (0.51, 1.3) | 0.44 |
| **Comorbidities** | | | | | | N= |
| None | Reference | | Reference | | Reference | |
| 1 comorbid condition | 1.03 (0.76, 1.4) | 0.84 | 1.10 (0.74, 1.6) | 0.63 | 0.94 (0.65, 1.4) | 0.75 |
| 2+ comorbid conditions | 0.81 (0.47, 1.4) | 0.44 | 1.48 (0.75, 2.9) | 0.26 | **0.47 (0.23, 1.0)** | **0.04** |

OR : odds ratio

[1]Adjusted for age and sex

[2]Adjusted for age

Table S9. Limited Analyses to Bus Santé participants

| | Overall sample[1] | | Women[2] | | Men[2] | |
|---|---|---|---|---|---|---|
| | OR (95% CI) | p-value | OR (95% CI) | p-value | OR (95% CI) | p-value |
| **Employment** | | | | | | |
| Employed | [Reference] | | [Reference] | | [Reference] | |
| Unemployed | 0.74 (0.43 – 1.3) | 0.27 | 0.71 (0.36 – 1.4) | 0.31 | 0.83 (0.33 – 2.1) | 0.70 |
| Student | 0.96 (0.42 – 2.2) | 0.92 | 1.3 (0.49 – 3.5) | 0.58 | 0.57 (0.13 – 2.5) | 0.46 |
| Retired | 0.60 (0.38 – 0.94) | 0.03 | 0.56 (0.29 – 1.1) | 0.08 | 0.65 (0.34 – 1.2) | 0.18 |
| Freelance/other | 1.1 (0.74 – 1.6) | 0.68 | 1.5 (0.87 – 2.5) | 0.15 | 0.82 (0.47 – 1.4) | 0.47 |
| **Occupational position** | | | | | | |
| Professional/Manager | [Reference] | | [Reference] | | [Reference] | |
| Higher grade WCW | 0.79 (0.50 – 1.2) | 0.29 | 1.1 (0.56 – 2.3) | 0.72 | 0.56 (0.30 – 1.1) | 0.08 |
| Independent | 0.46 (0.16 – 1.3) | 0.15 | 0.73 (0.16 – 3.4) | 0.68 | 0.34 (0.08 – 1.5) | 0.15 |
| Lower grade WCW | 1.1 (0.69 – 1.7) | 0.71 | 1.3 (0.64 – 2.6) | 0.49 | 1.0 (0.55 – 2.0) | 0.91 |
| BCW | 0.77 (0.41 – 1.5) | 0.42 | 0.43 (0.09 – 2.0) | 0.28 | 0.86 (0.42 – 1.8) | 0.68 |
| Other or N/A | 0.95 (0.61 – 1.5) | 0.82 | 1.3 (0.64 – 2.6) | 0.48 | 0.76 (0.42 – 1.4) | 0.37 |
| **Educational level** | | | | | | |
| Doctoral Education | [Reference] | | [Reference] | | [Reference] | |
| Haute Ecole/ University | 1.1 (0.64 – 2.0) | 0.67 | 1.8 (0.65 – 5.0) | 0.26 | 0.87 (0.43 – 1.7) | 0.69 |
| Secondary education | 1.0 (0.53 – 1.9) | 1.00 | 1.7 (0.57 – 5.0) | 0.35 | 0.71 (0.31 – 1.7) | 0.43 |
| Apprenticeship | 0.93 (0.49 – 1.8) | 0.83 | 1.2 (0.37 – 3.6) | 0.80 | 0.87 (0.40 – 1.9) | 0.74 |
| Mandatory education only | 1.2 (0.56 – 2.6) | 0.64 | 1.8 (0.51 – 6.3) | 0.37 | 1.0 (0.37 – 2.8) | 0.99 |
| Other | 1.2 (0.54 – 2.5) | 0.70 | 1.9 (0.55 – 6.5) | 0.31 | 0.87 (0.31 – 2.5) | 0.80 |
| **Area level income** | | | | | | |
| <37000 | [Reference] | | [Reference] | | [Reference] | |
| >=37000& <68000 | 0.86 (0.45 – 1.7) | 0.66 | 1.7 (0.53 – 4.4) | 0.30 | 0.55 (0.25 – 1.2) | 0.15 |
| >=68000 | 0.97 (0.35 – 2.7) | 0.95 | 1.7 (0.41 – 6.7) | 0.47 | 0.68 (0.18 – 2.5) | 0.55 |
| **Individual income** | | | | | | |
| >13000 | [Reference] | | [Reference] | | [Reference] | |
| 9500 – 13000 | 0.65 (0.44 – 0.95) | 0.03 | 0.40 (0.21 – 0.77) | 0.006 | 0.90 (0.56 – 1.4) | 0.65 |
| 7000 – 9499 | 0.56 (0.35 – 0.85) | 0.006 | 0.60 (0.27 – 0.92) | 0.03 | 0.64 (0.36 – 1.1) | 0.12 |
| 5000 – 6999 | 0.88 (0.58 – 1.3) | 0.54 | 1.0 (0.60 – 1.8) | 0.91 | 0.68 (0.34 – 1.3) | 0.26 |
| 3000 - 4999 | 0.83 (0.51 – 1.4) | 0.47 | 0.98 (0.52 – 1.8) | 0.94 | 0.61 (0.25 – 1.5) | 0.27 |
| <2999 CHF | 0.32 (0.11 – 0.89) | 0.03 | 0.47 (0.14 – 1.6) | 0.22 | 0.17 (0.02 – 1.3) | 0.08 |
| Don't Know/Refuse | 0.80 (0.55 – 1.2) | 0.26 | 1.0 (0.60 – 1.7) | 0.95 | 0.60 (0.34 – 1.1) | 0.08 |

[1]Adjusted for age and sex
[2]Adjusted for age

Table S10. Association between socio-demographic factors and IFA serological status in the SEROCOV-POP study population

| | Overall sample[1] | | Women[2] | | Men[2] | |
|---|---|---|---|---|---|---|
| | OR (95% CI) | P-value | OR (95% CI) | P-value | OR (95% CI) | P-value |
| **Employment** | | | | | | |
| Employed | [Reference] | | [Reference] | | [Reference] | |
| Unemployed | 0.68 (0.32 – 1.4) | 0.30 | 2.3 (0.56 – 9.8) | 0.24 | 0.03 (0.00 – 26.1) | 0.31 |
| Student | 0.75 (0.38 – 1.5) | 0.43 | 1.8 (0.42 – 7.8) | 0.42 | 0.10 (0.01 – 0.77) | 0.03 |
| Retired | 0.64 (0.31 – 1.3) | 0.23 | 0.90 (0.13 – 6.1) | 0.92 | 0.45 (0.06 – 3.6) | 0.45 |
| Freelance/other | 1.1 (0.61 – 1.9) | 0.82 | 2.1 (0.49 – 9.3) | 0.31 | 0.68 (0.13 – 3.5) | 0.65 |
| **Occupational position** | | | | | | |
| Professional/Manager | [Reference] | | [Reference] | | [Reference] | |
| Higher grade WCW | 0.75 (0.38 – 1.5) | 0.40 | 1.2 (0.24 – 6.2) | 0.80 | 0.51 (0.11 – 2.3) | 0.38 |
| Independent | 0.49 (0.12 – 2.0) | 0.32 | 2.8 (0.27 – 29.4) | 0.39 | 0.20 (0.00 – 9.3) | 0.41 |
| Lower grade WCW | 0.98 (0.49 – 2.0) | 0.96 | 2.1 (0.45 – 10.1) | 0.34 | 0.56 (0.09 – 3.6) | 0.54 |
| BCW | 0.40 (0.15 – 1.1) | 0.08 | 0.48 (0.04 – 6.4) | 0.58 | 0.53 (0.08 – 3.7) | 0.52 |
| Other or N/A | 0.79 (0.39 – 1.6) | 0.51 | 1.4 (0.27 – 7.4) | 0.67 | 0.10 (0.01 – 1.3) | 0.08 |
| **Educational level** | | | | | | |
| Doctoral Education | [Reference] | | [Reference] | | [Reference] | |
| Haute Ecole/ University | 1.4 (0.77 - 2.7) | 0.32 | 2.6 (1.1 – 8.7) | 0.06 | 1.4 (0.77 – 2.7) | 0.32 |
| Secondary education | 1.4 (0.71 – 2.7) | 0.39 | 2.8 (1.1 – 9.4) | 0.05 | 1.4 (0.71 – 2.7) | 0.39 |
| Apprenticeship | 1.1 (0.57 – 2.2) | 0.80 | 2.4 (0.93 – 8.2) | 0.11 | 1.1 (0.57 – 2.2) | 0.80 |
| Mandatory education only | 0.81 (0.33 – 1.9) | 0.63 | 2.1 (0.73 – 7.5) | 0.20 | 0.81 (0.33 – 1.9) | 0.63 |
| Other | 1.1 (0.44 – 2.7) | 0.83 | 3.3 (1.1 – 11.8) | 0.04 | 1.1 (0.44 – 2.7) | 0.83 |
| **Area level income** | | | | | | |
| <37000 | [Reference] | | [Reference] | | [Reference] | |
| >=37000& <68000 | 1.2 (0.41 - 3.8) | 0.70 | 1.5 (0.69 – 3.4) | 0.30 | 0.99 (0.49- 2.0) | 0.98 |
| >=68000 | 1.4 (0.26 – 7.1) | 0.72 | 1.7 (0.55- 5.3) | 0.36 | 1.1 (0.36 – 3.2) | 0.91 |
| **Individual income** | | | | | | |
| >13000 | [Reference] | | [Reference] | | [Reference] | |
| 9500 – 13000 | 0.75 (0.51 – 1.1) | 0.16 | 0.61 (0.31 – 1.2) | 0.14 | 0.90 (0.55 – 1.5) | 0.66 |
| 7000 – 9499 | 0.56 (0.36 – 0.88) | 0.01 | 0.61 (0.32 – 1.2) | 0.14 | 0.56 (0.30 – 1.0) | 0.06 |
| 5000 – 6999 | 1.1 (0.69 – 1.6) | 0.82 | 1.4 (0.80 – 2.5) | 0.23 | 0.73 (0.37 – 1.4) | 0.36 |
| 3000 - 4999 | 0.92 (0.55 – 1.5) | 0.76 | 1.5 (0.78 – 2.8) | 0.24 | 0.32 (0.10 – 1.1) | 0.06 |
| <2999 CHF | 0.27 (0.08 – 0.87) | 0.03 | 0.39 (0.09 – 1.7) | 0.21 | 0.18 (0.02 – 1.3) | 0.09 |
| Don't Know/Refuse | 0.79 (0.53 – 1.2) | 0.26 | 1.1 (0.59 – 1.9) | 0.84 | 0.61 (0.34 – 1.1) | 0.10 |

[1]Adjusted for age and sex

[2]Adjusted for age

### Section 3: Symptoms

Table S11. Uni- and multivariate Odds Ratios (OR) of IgG seropositivity (IgG ≥ 1.1) according to each symptom, by age category

| **Symptom** | **Overall sample, OR (95% CI)** | | **< 10 years, OR (95% CI)** | | **10-17 years, OR (95% CI)** | | **18-49 years, OR (95% CI)** | | **50-64 years, OR (95% CI)** | | **65-74 years, OR (95% CI)** | | **> 74 years, OR (95% CI)** | |
|---|---|---|---|---|---|---|---|---|---|---|---|---|---|---|
| | univariate OR | adjusted OR[1] | univariate OR | adjusted OR[2] | univariate OR | adjusted OR[2] | univariate OR | adjusted OR[2] | univariate OR | adjusted OR[2] | univariate OR | adjusted OR[2] | univariate OR | adjusted OR[2] |
| Fever | 4.9(4.2-5.9) | 2.2(1.7-2.8) | 1.6(0.38-6.1) | 0.94(0.13-5.6) | 1.8(0.92-3.4) | 1.6(0.63-3.9) | 4.2(3.3-5.5) | 2.2(1.5-3.1) | 7.8(5.7-10.8) | 2.2(1.4-3.4) | 5.3(2.9-9.4) | 2.8(1.2-6.4) | 9.9(3.5-27.4) | 19.6(3.8-116.1) |
| Cough | 2.8(2.3-3.3) | 1.1(0.89-1.5) | 2.1(0.52-8.3) | 2.1(0.33-13.1) | 1.5(0.78-2.8) | 0.74(0.27-2) | 2.1(1.6-2.7) | 1(0.7-1.5) | 4.5(3.3-6.2) | 1.6(1-2.5) | 2.6(1.4-4.5) | 0.87(0.36-2) | 4.4(1.6-12) | 0.59(0.09-3.1) |
| Rhinorrhea / sneezing | 2(1.7-2.3) | 0.86(0.68-1.08) | 1.3(0.32-5.2) | 0.63(0.08-3.7) | 1.1(0.61-2) | 0.66(0.27-1.5) | 1.7(1.3-2.2) | 0.79(0.56-1.1) | 2.5(1.8-3.4) | 1(0.66-1.5) | 2.1(1.2-3.6) | 0.96(0.45-2) | 2.2(0.74-5.9) | 1.5(0.37-5.3) |
| Sore throat | 1.7(1.4-2) | 0.63(0.49-0.81) | 1.6(0.33-6.3) | 0.91(0.11-6.9) | 1.4(0.72-2.7) | 1.5(0.61-3.7) | 1.3(1-1.7) | 0.61(0.42-0.88) | 2(1.5-2.7) | 0.52(0.33-0.81) | 2.5(1.3-4.4) | 0.74(0.29-1.8) | 2(0.44-6.4) | 0.96(0.13-5.2) |
| Dyspnea | 3.5(2.8-4.3) | 0.93(0.68-1.3) | 0 (NA) | 0 (NA) | 1(0.16-3.6) | 0.44(0.05-2.4) | 3.1(2.3-4.2) | 0.98(0.63-1.5) | 4.1(2.8-5.9) | 0.8(0.45-1.4) | 4.3(2.1-8.3) | 1.3(0.45-3.3) | 5.1(1.5-14.8) | 2(0.25-12.9) |
| Headache | 3.3(2.7-3.9) | 1.5(1.2-1.9) | 1(0.15-4.5) | 0.51(0.05-3.6) | 1.1(0.55-2.1) | 0.72(0.29-1.7) | 2.7(2.1-3.5) | 1.6(1.1-2.3) | 5.2(3.8-7.1) | 1.9(1.3-3) | 2.9(1.6-5.2) | 1.1(0.47-2.6) | 2.4(0.53-7.7) | 0.32(0.03-2.2) |
| Myalgia / arthralgia | 4.2(3.5-5) | 1.6(1.2-2) | 2.6(0.38-11.7) | 1.9(0.17-15.2) | 1.6(0.62-3.4) | 1(0.32-3) | 4.7(3.6-6) | 1.7(1.2-2.5) | 5.2(3.8-7.1) | 1.6(1-2.5) | 2.3(1.2-4.2) | 0.92(0.38-2.1) | 2(0.53-5.7) | 0.94(0.13-4.9) |
| Fatigue | 4.3(3.6-5.1) | 1.6(1.2-2) | 2.3(0.47-9.1) | 2.7(0.28-21.9) | 2(1-3.8) | 1.7(0.65-4.4) | 3.7(2.9-4.8) | 1.3(0.89-1.9) | 5.4(4-7.5) | 1.9(1.2-3) | 5.4(3-9.4) | 2.9(1.2-6.7) | 4(1.4-10.9) | 0.97(0.16-4.8) |
| Loss of appetite | 6.2(4.9-7.8) | 1.2(0.83-1.6) | 1.3(0.07-7.8) | 1.1(0.04-13.2) | 1.4(0.32-4.1) | 0.41(0.06-2) | 5.5(3.9-7.7) | 1.4(0.85-2.3) | 10.7(7.2-15.8) | 1.4(0.76-2.4) | 5.3(2.4-10.8) | 0.48(0.13-1.7) | 9.1(2.7-27.6) | 10.1(1.4-80.5) |
| Nausea / vomiting | 2(1.5-2.7) | 0.47(0.31-0.71) | 0 (NA) | 0 (NA) | 1.3(0.39-3.6) | 0.59(0.12-2.3) | 1.5(0.92-2.3) | 0.54(0.28-1) | 2.8(1.6-4.5) | 0.51(0.23-1.1) | 3.3(1.1-8) | 0.82(0.16-3.8) | 3.8(0.56-15.2) | 0.26(0.01-3.7) |
| Diarrhea | 2.4(1.9-3) | 1.1(0.8-1.4) | 2.9(0.42-12.9) | 2.9(0.3-18.5) | 2.9(1.3-5.9) | 3.3(1.2-8.3) | 1.7(1.2-2.3) | 0.88(0.56-1.4) | 3.4(2.4-4.9) | 1.3(0.76-2.1) | 2(0.86-4.2) | 0.61(0.18-1.7) | 2.4(0.36-9.1) | 0.57(0.03-5.5) |
| Abdominal pain | 1.7(1.3-2.2) | 0.53(0.35-0.77) | 2.3(0.33-10.2) | 1.6(0.13-11.2) | 1.1(0.33-3) | 0.53(0.11-2) | 1.5(1-2.1) | 0.71(0.4-1.2) | 1.9(1.1-3.1) | 0.39(0.18-0.8) | 2.4(0.9-5.5) | 0.68(0.16-2.4) | 1.2(0.06-6.1) | 0.06(0-0.92) |
| Anosmia / dysgeusia | 24.8(20.1-30.6) | 16.8(13.1-21.8) | 0 (NA) | 0 (NA) | 16.1(6.8-38.2) | 24.3(8.8-72.6) | 32.6(23.8-45) | 22.5(15.6-32.7) | 28.1(19.3-40.9) | 14.4(9-23.3) | 11.7(6.1-21.9) | 9(3.6-22.4) | 6.1(1.3-21.4) | 3.5(0.28-37.7) |
| Asymptomatic | 0.2(0.16-0.26) | 0.21(0.16-0.26) | 1.4(0.34-5.4) | 1.6(0.38-6.2) | 0.43(0.22-0.81) | 0.43(0.22-0.8) | 0.21(0.14-0.31) | 0.21(0.14-0.3) | 0.13(0.07-0.21) | 0.12(0.07-0.2) | 0.24(0.12-0.45) | 0.24(0.12-0.44) | 0.18(0.05-0.53) | 0.17(0.05-0.48) |

[1]Adjusted for age, sex and all symptoms

[2]Adjusted for sex and all symptoms

Table S12. Symptoms by age group classified by SARS-CoV-2 IgG serological status (positive result: IgG ≥ 1.1)

| **Symptom** | **< 10 years** | | | **10-17 years** | | | **18-49 years** | | | **50-64 years** | | | **65-74 years** | | | **> 74 years** | | |
|---|---|---|---|---|---|---|---|---|---|---|---|---|---|---|---|---|---|---|
| | Negative N=265 | Positive N = 9 | p-value | Negative N=578 | Positive N=50 | p-value | Negative N=2,830 | Positive N=278 | p-value | Negative N=2,514 | Positive N=180 | p-value | Negative N=1,140 | Positive N=56 | p-value | Negative N=427 | Positive N=17 | p-value |
| Asymptomatic | 90 (37%) | 4 (44%) | 0.7 | 252 (45%) | **13 (26%)** | **0.015** | 976 (35%) | **28 (10%)** | **<0.001** | 1,092 (44%) | **16 (8.9%)** | **<0.001** | 604 (54%) | **12 (22%)** | **<0.001** | 258 (63%) | **4 (24%)** | **0.003** |
| Fever | 83 (34%) | 4 (44%) | 0.5 | 108 (19%) | 15 (30%) | 0.1 | 484 (17%) | **130 (47%)** | **<0.001** | 315 (13%) | **95 (53%)** | **<0.001** | 112 (10%) | **20 (37%)** | **<0.001** | 34 (8.3%) | **8 (47%)** | **<0.001** |
| Cough | 67 (27%) | 4 (44%) | 0.3 | 134 (24%) | 16 (32%) | 0.3 | 778 (28%) | **123 (44%)** | **<0.001** | 546 (22%) | **100 (56%)** | **<0.001** | 209 (19%) | **20 (37%)** | **0.002** | 56 (14%) | **7 (41%)** | **0.006** |
| Sneezing / Rhinorrhea | 92 (37%) | 4 (44%) | 0.7 | 198 (35%) | 19 (38%) | 0.8 | 1,064 (38%) | **141 (51%)** | **<0.001** | 720 (29%) | **90 (50%)** | **<0.001** | 312 (28%) | **24 (44%)** | **0.013** | 82 (20%) | 6 (35%) | 0.13 |
| Sore throat | 58 (24%) | 3 (33%) | 0.5 | 120 (21%) | 14 (28%) | 0.4 | 830 (29%) | 97 (35%) | 0.071 | 563 (23%) | **66 (37%)** | **<0.001** | 175 (16%) | **17 (31%)** | **0.004** | 40 (9.7%) | 3 (18%) | 0.2 |
| Dyspnea | 8 (3.3%) | 0 (0%) | >0.9 | 22 (3.9%) | 2 (4.0%) | >0.9 | 267 (9.5%) | **69 (25%)** | **<0.001** | 179 (7.2%) | **43 (24%)** | **<0.001** | 70 (6.3%) | **12 (22%)** | **<0.001** | 31 (7.5%) | **5 (29%)** | **0.009** |
| Headache | 53 (22%) | 2 (22%) | >0.9 | 136 (24%) | 13 (26%) | >0.9 | 927 (33%) | **159 (57%)** | **<0.001** | 634 (25%) | **114 (64%)** | **<0.001** | 152 (14%) | **17 (31%)** | **<0.001** | 34 (8.3%) | 3 (18%) | 0.2 |
| Myalgia / arthralgia | 24 (9.8%) | 2 (22%) | 0.2 | 53 (9.4%) | 7 (14%) | 0.3 | 472 (17%) | **135 (49%)** | **<0.001** | 405 (16%) | **90 (50%)** | **<0.001** | 150 (13%) | **14 (26%)** | **0.017** | 56 (14%) | 4 (24%) | 0.3 |
| Fatigue | 44 (18%) | 3 (33%) | 0.4 | 91 (16%) | 14 (28%) | 0.055 | 850 (30%) | **171 (62%)** | **<0.001** | 586 (23%) | **112 (63%)** | **<0.001** | 155 (14%) | **25 (46%)** | **<0.001** | 61 (15%) | **7 (41%)** | **0.01** |
| Loss of appetite | 21 (8.5%) | 1 (11%) | 0.6 | 25 (4.5%) | 3 (6.0%) | 0.5 | 126 (4.5%) | **57 (21%)** | **<0.001** | 87 (3.5%) | **50 (28%)** | **<0.001** | 46 (4.1%) | **10 (19%)** | **<0.001** | 18 (4.4%) | **5 (29%)** | **0.001** |
| Nausea | 14 (5.7%) | 0 (0%) | >0.9 | 34 (6.1%) | 4 (8.0%) | 0.5 | 170 (6.0%) | 24 (8.6%) | 0.12 | 108 (4.3%) | **20 (11%)** | **<0.001** | 34 (3.0%) | **5 (9.3%)** | **0.03** | 14 (3.4%) | 2 (12%) | 0.13 |
| Diarrhea | 22 (8.9%) | 2 (22%) | 0.2 | 45 (8.0%) | **10 (20%)** | **0.009** | 342 (12%) | **53 (19%)** | **0.001** | 229 (9.2%) | **46 (26%)** | **<0.001** | 89 (8.0%) | 8 (15%) | 0.079 | 22 (5.4%) | 2 (12%) | 0.2 |
| Abdominal pain | 27 (11%) | 2 (22%) | 0.3 | 40 (7.1%) | 4 (8.0%) | 0.8 | 238 (8.5%) | 33 (12%) | 0.071 | 146 (5.8%) | **19 (11%)** | **0.016** | 55 (4.9%) | 6 (11%) | 0.056 | 21 (5.1%) | 1 (5.9%) | 0.6 |
| Anosmia/ dysgeusia | 2 (0.8%) | 0 (0%) | >0.9 | 12 (2.1%) | **13 (26%)** | **<0.001** | 86 (3.1%) | **141 (51%)** | **<0.001** | 73 (2.9%) | **82 (46%)** | **<0.001** | 46 (4.1%) | **18 (33%)** | **<0.001** | 14 (3.4%) | **3 (18%)** | **0.025** |
| Other symptoms | 2 (0.8%) | 1 (11%) | 0.1 | 4 (0.7%) | 2 (4.0%) | 0.08 | 61 (2.2%) | 6 (2.2%) | >0.9 | 72 (2.9%) | 9 (5.0%) | 0.2 | 29 (2.6%) | 0 (0%) | 0.6 | 13 (3.2%) | 0 (0%) | >0.9 |

Table S13. Frequency of symptoms reported in the overall sample according to IFA results

| Symptom | Overall (N = 8,344) | SARS-CoV-2 IgG test result<br>Negative (N = 7,813)* | <br>Positive (N = 531) | p-value |
|---|---|---|---|---|
| Fatigue | 2,119 (26%) | 1,795 (23%) | 324 (61%) | <0.001 |
| Headache | 2,244 (27%) | 1,945 (25%) | 299 (56%) | <0.001 |
| Sneezing/Rhinorrhea | 2,752 (33%) | 2,484 (32%) | 268 (50%) | <0.001 |
| Fever | 1,408 (17%) | 1,141 (15%) | 267 (50%) | <0.001 |
| Cough | 2,060 (25%) | 1,800 (23%) | 260 (49%) | <0.001 |
| Anosmia / dysgeusia | 490 (6.0%) | 215 (2.8%) | 275 (52%) | <0.001 |
| Myalgia / arthralgia | 1,412 (17%) | 1,169 (15%) | 243 (46%) | <0.001 |
| Sore throat | 1,986 (24%) | 1,810 (23%) | 176 (33%) | <0.001 |
| Dyspnea | 708 (8.6%) | 576 (7.5%) | 132 (25%) | <0.001 |
| Loss of appetite | 449 (5.5%) | 320 (4.2%) | 129 (24%) | <0.001 |
| Diarrhea | 870 (11%) | 752 (9.8%) | 118 (22%) | <0.001 |
| Abdominal pain | 592 (7.2%) | 531 (6.9%) | 61 (11%) | <0.001 |
| Nausea / vomiting | 429 (5.2%) | 370 (4.8%) | 59 (11%) | <0.001 |
| Other symptoms | 199 (2.4%) | 182 (2.4%) | 17 (3.2%) | 0.3 |
| Asymptomatic | 3,349 (41%) | 3,295 (43%) | 54 (10%) | <0.001 |

* 110 missing values not included

Table S14. Uni- and multivariate Odds Ratios (OR) of seropositivity by IFA according to each symptom, by age category

| Symptom | Overall sample, OR (95% CI) | | < 10 years, OR (95% CI) | | 10-17 years, OR (95% CI) | | 18-49 years, OR (95% CI) | | 50-64 years, OR (95% CI) | | 65-74 years, OR (95% CI) | | [illegible]4 years, OR (95% CI) | |
|---|---|---|---|---|---|---|---|---|---|---|---|---|---|---|
| | Univariate OR | Adjusted OR[1] | Univariate OR | Adjusted OR[2] | Univariate OR | Adjusted OR[2] | Univariate OR | Adjusted OR[2] | Univariate OR | Adjusted OR[2] | Univariate OR | Adjusted OR[2] | Univariate OR | Adjusted OR[2] |
| Fever | 5.8(4.9-7) | 2.5(1.9-3.2) | 2(0.36-10.8) | 1.3(0.13-12.1) | 2(0.98-3.8) | 1.4(0.5-3.7) | 5(3.8-6.5) | 2.6(1.8-3.9) | 9.3(6.7-13) | 2.6(1.6-4.1) | 6.2(3.2-11.5) | 2.8(1.1-7) | 11.1(3.9-32.1) | 27.9(4.8-193.6) |
| Cough | 3.1(2.6-3.8) | 1.2(0.93-1.6) | 2.7(0.48-14.7) | 3.1(0.31-32) | 2.1(1.1-3.9) | 1.4(0.5-3.9) | 2.3(1.7-2.9) | 0.99(0.65-1.5) | 5.1(3.7-7.1) | 1.7(1.1-2.8) | 2.7(1.4-5) | 0.82(0.3-2.1) | 4.9(1.7-13.8) | 0.69(0.1-4.1) |
| Rhinorrhea / sneezing | 2.1(1.8-2.6) | 0.85(0.66-1.1) | 1.7(0.31-9.2) | 0.79(0.07-7.4) | 1.2(0.6-2.2) | 0.51(0.19-1.3) | 1.9(1.5-2.5) | 0.8(0.54-1.2) | 2.7(2-3.7) | 1.1(0.73-1.8) | 1.9(1-3.6) | 0.56(0.22-1.3) | 1.8(0.56-5.1) | 1.1(0.23-4.3) |
| Sore throat | 1.6(1.3-1.9) | 0.46(0.34-0.6) | 1.6(0.22-8.5) | 0.58(0.04-6.2) | 1.2(0.57-2.4) | 0.78(0.27-2.1) | 1.3(1-1.8) | 0.54(0.36-0.8) | 1.7(1.2-2.3) | 0.28(0.17-0.47) | 2.5(1.3-4.7) | 0.5(0.17-1.4) | 1.3(0.2-4.8) | 0.33(0.02-2.6) |
| Dyspnea | 4.1(3.3-5.1) | 0.94(0.67-1.3) | 0(NA) | 0(NA) | 1.9(0.44-5.8) | 0.89(0.13-4.2) | 3.4(2.5-4.5) | 0.78(0.47-1.3) | 5.2(3.5-7.5) | 1.1(0.61-2) | 5(2.3-10) | 1.3(0.43-3.7) | 5.6(1.7-16.4) | 3.4(0.41-23.2) |
| Headache | 3.8(3.2-4.6) | 1.6(1.3-2.1) | 0.72(0.04-4.6) | 0.26(0.01-3.2) | 1.3(0.65-2.6) | 0.69(0.26-1.7) | 3.2(2.5-4.2) | 1.9(1.3-2.8) | 5.6(4-7.8) | 1.9(1.2-3.1) | 4(2.1-7.5) | 1.7(0.65-4.1) | 2.6(0.57-8.4) | 0.22(0.01-1.9) |
| Myalgia / arthralgia | 4.7(3.9-5.7) | 1.5(1.2-2) | 1.8(0.09-11.7) | 1.2(0.03-20.8) | 1.8(0.72-4.1) | 1(0.28-3.2) | 5(3.9-6.6) | 1.4(0.91-2.1) | 5.8(4.2-8) | 1.8(1.1-2.9) | 3(1.5-5.8) | 1.2(0.48-3) | 2.1(0.58-6.3) | 1.3(0.16-6.9) |
| Fatigue | 5.2(4.3-6.2) | 1.7(1.3-2.3) | 2.3(0.31-12) | 4.6(0.25-69.3) | 2.7(1.4-5.2) | 2.5(0.87-6.7) | 4.5(3.4-5.9) | 1.4(0.93-2.2) | 6(4.3-8.4) | 1.8(1.1-3) | 8.3(4.5-15.6) | 5(1.9-12.8) | 3.4(1.1-9.5) | 0.41(0.05-2.4) |
| Loss of appetite | 7.4(5.9-9.3) | 1.2(0.82-1.7) | 0(NA) | 0(NA) | 2.3(0.64-6.2) | 0.39(0.06-2) | 6.5(4.6-9.1) | 1.5(0.84-2.5) | 12.8(8.6-18.9) | 1.4(0.75-2.5) | 6.9(3.1-14.4) | 0.49(0.13-1.7) | 9.9(2.9-30.7) | 16.8(1.8-195.8) |
| Nausea / vomiting | 2.5(1.8-3.3) | 0.54(0.35-0.83) | 0(NA) | 0(NA) | 2.1(0.68-5.2) | 0.63(0.13-2.5) | 1.9(1.2-2.9) | 0.72(0.37-1.3) | 3(1.8-4.9) | 0.45(0.19-1) | 4.1(1.4-10.3) | 2.4(0.45-10.9) | 4.1(0.6-16.5) | 0.25(0.01-4) |
| Diarrhea | 2.6(2.1-3.3) | 1.1(0.8-1.5) | 5.2(0.69-28) | 9.9(0.82-103.4) | 3.4(1.5-7.1) | 3.3(1.1-8.7) | 1.8(1.3-2.5) | 0.8(0.48-1.3) | 3.8(2.6-5.4) | 1.4(0.79-2.3) | 2.2(0.87-4.8) | 0.68(0.19-2.1) | 2.5(0.38-9.8) | 0.45(0.02-5) |
| Stomach ache | 1.8(1.3-2.3) | 0.42(0.27-0.65) | 1.6(0.08-10.3) | 0.98(0.03-13.7) | 1.7(0.57-4.3) | 0.89(0.19-3.3) | 1.6(1.1-2.4) | 0.74(0.4-1.3) | 1.9(1.1-3.1) | 0.28(0.12-0.64) | 1.4(0.32-3.9) | 0.09(0.01-0.53) | 1.2(0.07-6.6) | 0.07(0-1) |
| Anosmia / dysgeusia | 37.4(30.1-46.6) | 26.4(20.2-34.6) | 0(NA) | 0(NA) | 23.6(9.9-57.6) | 25.8(9.2-77.8) | 55.5(39.6-78.4) | 40.4(27.3-60.7) | 40.1(27.3-59.3) | 23.7(14.3-39.9) | 16.3(8.3-31.7) | 14.2(5.4-38) | 6.6(1.4-23.3) | 4.2(0.28-53.8) |
| Asymptomatic | 0.15(0.11-0.2) | 0.16(0.12-0.21) | 0.85(0.12-4.5) | 0.85(0.12-4.6) | 0.31(0.14-0.63) | 0.31(0.14-0.63) | 0.14(0.08-0.22) | 0.13(0.08-0.21) | 0.09(0.05-0.16) | 0.09(0.04-0.16) | 0.25(0.12-0.5) | 0.25(0.11-0.48) | 0.27(0.09-0.77) | 0.25(0.08-0.72) |
| Close contact | 5.8(4.9-7) | 2.5(1.9-3.2) | 2(0.36-10.8) | 1.3(0.13-12.1) | 2(0.98-3.8) | 1.4(0.5-3.7) | 5(3.8-6.5) | 2.6(1.8-3.9) | 9.3(6.7-13) | 2.6(1.6-4.1) | 6.2(3.2-11.5) | 2.8(1.1-7) | 11.1(3.9-32.1) | 27.9(4.8-193.6) |

[1]Adjusted for age, sex and all symptoms

[2]Adjusted for sex and all symptoms

Table S15. Frequency of symptoms reported in the overall sample for participants with a unique symptomatic episode (positive result: IgG ≥ 1.1)

| Symptom | Overall (N = 6,700) | SARS-CoV-2 IgG test result<br>Negative (N = 6,229)* | <br>Positive (N = 471)** | p-value |
|---|---|---|---|---|
| Fatigue | 1,349 (20%) | 1,097 (18%) | 252 (54%) | <0.001 |
| Headache | 1,310 (20%) | 1,084 (18%) | 226 (48%) | <0.001 |
| Sneezing/Rhinorrhea | 1,702 (26%) | 1,503 (25%) | 199 (43%) | <0.001 |
| Fever | 1,032 (16%) | 812 (13%) | 220 (47%) | <0.001 |
| Cough | 1,413 (21%) | 1,209 (20%) | 204 (44%) | <0.001 |
| Anosmia / dysgeusia | 383 (5.8%) | 173 (2.8%) | 210 (45%) | <0.001 |
| Myalgia / arthralgia | 868 (13%) | 682 (11%) | 186 (40%) | <0.001 |
| Sore throat | 1,222 (19%) | 1,088 (18%) | 134 (29%) | <0.001 |
| Dyspnea | 463 (7.0%) | 362 (5.9%) | 101 (22%) | <0.001 |
| Loss of appetite | 319 (4.8%) | 221 (3.6%) | 98 (21%) | <0.001 |
| Diarrhea | 470 (7.1%) | 379 (6.2%) | 91 (19%) | <0.001 |
| Abdominal pain | 295 (4.5%) | 251 (4.1%) | 44 (9.4%) | <0.001 |
| Nausea / vomiting | 229 (3.5%) | 189 (3.1%) | 40 (8.5%) | <0.001 |
| Other symptoms | 144 (2.2%) | 127 (2.1%) | 17 (3.6%) | 0.04 |
| Asymptomatic | 3,346 (51%) | 3,269 (53%) | 77 (16%) | <0.001 |

* 107 missing values not included ** 3 missing values not included

Table S16. Uni- and multivariate Odds Ratios (OR) of seropositivity according to each symptom, by age category, in participants with a unique symptomatic episode (positive result: IgG ≥ 1.1)

| **Symptom** | **Overall sample, OR (95% CI)** | | **< 10 years, OR (95% CI)** | | **10-17 years, OR (95% CI)** | | **18-49 years, OR (95% CI)** | | **50-64 years, OR (95% CI)** | | **65-74 years, OR (95% CI)** | | **> 74 years, OR (95% CI)** | |
|---|---|---|---|---|---|---|---|---|---|---|---|---|---|---|
| | Univariate OR | Adjusted OR[1] | Univariate OR | Adjusted OR[2] | Univariate OR | Adjusted OR[2] | Univariate OR | Adjusted OR[2] | Univariate OR | Adjusted OR[2] | Univariate OR | Adjusted OR[2] | Univariate OR | Adjusted OR[2] |
| Fever | 5.8(4.8-7.1) | 2.5(1.9-3.3) | 0.51(0.03-3.3) | 1.5(0.07-10.9) | 1.6(0.75-3.2) | 1.4(0.47-4) | 5.3(4-7.1) | 2.5(1.7-3.9) | 9.6(6.7-14) | 2.7(1.6-4.6) | 5.8(3.1-10.9) | 3.3(1.3-8.3) | 10.4(3.7-29.3) | 16.7(3-106.1) |
| Cough | 3.1(2.6-3.8) | 1.1(0.8-1.5) | 0.71(0.04-4.5) | 1.5(0.07-12.5) | 1.8(0.87-3.5) | 1.3(0.4-3.8) | 2.5(1.9-3.3) | 1(0.64-1.6) | 5.1(3.5-7.3) | 1.4(0.82-2.5) | 2.6(1.4-4.8) | 0.43(0.13-1.2) | 5.6(1.9-15.4) | 0.61(0.07-3.9) |
| Rhinorrhea / sneezing | 2.3(1.9-2.8) | 0.82(0.62-1.1) | 0.49(0.03-3.1) | 0.92(0.04-7) | 1.1(0.55-2.2) | 0.66(0.23-1.7) | 2(1.5-2.7) | 0.82(0.53-1.2) | 2.8(1.9-4) | 0.86(0.5-1.5) | 2.5(1.3-4.5) | 1.07(0.42-2.6) | 3.1(1-8.5) | 2.3(0.49-9.1) |
| Sore throat | 1.9(1.5-2.3) | 0.5(0.36-0.68) | 0(NA) | 0(NA) | 1.1(0.45-2.3) | 0.87(0.26-2.6) | 1.3(1-1.8) | 0.43(0.27-0.7) | 2.7(1.8-3.9) | 0.56(0.32-0.97) | 2.5(1.2-4.8) | 0.54(0.17-1.6) | 3.1(0.68-10.3) | 0.67(0.06-5.3) |
| Dyspnea | 4.4(3.4-5.6) | 0.93(0.63-1.3) | 0(NA) | 3.7(NA) | 2.4(0.36-9.8) | 2.5(0.2-20.1) | 3.9(2.8-5.5) | 0.94(0.55-1.6) | 4.8(3-7.5) | 0.63(0.3-1.3) | 5.4(2.4-11.3) | 2.1(0.58-7.3) | 4.9(1.5-14.2) | 0.95(0.08-8) |
| Headache | 4.3(3.6-5.3) | 1.6(1.2-2.2) | 0(NA) | 0(NA) | 1.3(0.55-2.6) | 0.79(0.25-2.3) | 3.9(3-5.2) | 1.9(1.2-2.9) | 7(4.9-10.2) | 2(1.1-3.5) | 3.5(1.7-6.8) | 0.94(0.31-2.6) | 3.1(0.68-10.3) | 0.32(0.02-2.4) |
| Myalgia / arthralgia | 5.3(4.3-6.4) | 1.4(1-1.9) | 0(NA) | 0(NA) | 1(0.23-2.9) | 0.47(0.07-2.2) | 6(4.5-8) | 1.4(0.9-2.2) | 7.2(4.9-10.4) | 1.7(1-2.9) | 2.9(1.4-5.7) | 0.83(0.28-2.3) | 2.7(0.73-8) | 0.7(0.07-5) |
| Fatigue | 5.3(4.4-6.5) | 1.7(1.3-2.3) | 0(NA) | 0(NA) | 1.9(0.86-4) | 1.9(0.59-5.7) | 5.2(3.9-7) | 1.6(0.99-2.5) | 6.3(4.4-9.1) | 1.7(0.96-3.1) | 7.1(3.9-13.2) | 4.7(1.7-12.3) | 5(1.8-13.8) | 1.72(0.26-9.2) |
| Loss of appetite | 7.1(5.4-9.1) | 1.2(0.8-1.8) | 0(NA) | 0(NA) | 0.61(0.03-3.1) | 0.65(0.03-4.3) | 7(4.7-10.4) | 1.2(0.66-2.2) | 11.2(7.1-17.6) | 1.7(0.82-3.3) | 6.1(2.6-13.2) | 0.67(0.15-2.6) | 12.4(3.5-39.8) | 21.5(2.2-249.7) |
| Nausea / vomiting | 2.9(2-4.1) | 0.46(0.27-0.78) | 0(NA) | 0(NA) | 1.7(0.39-5.3) | 0.9(0.13-4.3) | 2.6(1.4-4.3) | 0.57(0.25-1.2) | 3.4(1.7-6.3) | 0.28(0.1-0.75) | 4.6(1.5-11.7) | 1.5(0.23-9.3) | 5.3(0.77-23.1) | 0.24(0.01-4.8) |
| Diarrhea | 3.7(2.8-4.7) | 1.2(0.82-1.7) | 0(NA) | 0(NA) | 2.6(0.83-6.7) | 2.1(0.44-7.7) | 3.1(2.1-4.5) | 1(0.57-1.8) | 5.2(3.3-7.9) | 1.7(0.86-3.1) | 3.1(1.31-6.7) | 0.79(0.21-2.5) | 3.4(0.5-13.6) | 0.42(0.02-5.4) |
| Abdominal pain | 2.4(1.7-3.4) | 0.42(0.25-0.69) | 0(NA) | 0(NA) | 0(NA) | 0(NA) | 2.7(1.7-4.2) | 0.77(0.37-1.6) | 2.9(1.5-5.2) | 0.39(0.14-1) | 2.4(0.69-6.3) | 0.19(0.02-1.2) | 1.7(0.09-9.5) | 0.19(0-3.8) |
| Anosmia / dysgeusia | 28(22.1-35.5) | 19.5(14.4-26.5) | 0(NA) | 0.12(NA) | 21.6(8.3-59) | 30.3(9.6-111.2) | 38.5(26.9-55.7) | 25.8(16.7-40.3) | 30.5(19.7-47.5) | 16(9-28.9) | 12.2(5.9-24.3) | 9.2(3.1-27.4) | 7(1.5-25.5) | 2.6(0.19-29.1) |
| Asymptomatic | 0.17(0.13-0.22) | 0.18(0.14-0.23) | 2.4(0.47-17.9) | 2.5(0.48-18.7) | 0.35(0.17-0.68) | 0.35(0.17-0.68) | 0.17(0.11-0.25) | 0.17(0.11-0.25) | 0.12(0.07-0.19) | 0.11(0.06-0.19) | 0.21(0.1-0.41) | 0.21(0.1-0.4) | 0.13(0.04-0.38) | 0.11(0.03-0.34) |
| Close contact | 5.8(4.8-7.1) | 2.5(1.9-3.3) | 0.51(0.03-3.3) | 1.5(0.07-10.9) | 1.6(0.75-3.2) | 1.4(0.47-4) | 5.3(4-7.1) | 2.5(1.7-3.9) | 9.6(6.7-14) | 2.7(1.6-4.6) | 5.8(3.1-10.9) | 3.3(1.3-8.3) | 10.4(3.7-29.3) | 16.7(3-106.1) |

[1]Adjusted for age, sex and all symptoms

[2]Adjusted for sex and all symptoms
